# Comorbidity clusters associated with newly treated Type 2 diabetes mellitus: a Bayesian nonparametric analysis

**DOI:** 10.1101/2022.04.07.22273569

**Authors:** Adrian Martinez-De la Torre, Fernando Perez-Cruz, Stefan Weiler, Andrea M. Burden

## Abstract

**Background:** Type 2 Diabetes Mellitus (T2DM) is associated with the development of chronic comorbidities over time, which can lead to high drug utilization and adverse events. Understanding the patterns of disease progression is needed.

**Objectives:** To identify common comorbidity clusters and explore the progression over time in newly treated T2DM patients.

**Methods:** The IQVIA Medical Research Data incorporating data from THIN, a Cegedim database of anonymized electronic health records, was used to identify all patients with a first-ever prescription for a non-insulin antidiabetic drug (NIAD) between January 2006 and December 2019. We selected 58 chronic comorbidities of interest and used Bayesian nonparametric latent models (BNLM) to identify disease clusters and model their progression over time.

**Results:** Among the 175,383 eligible T2DM patients, we identified the 20 most frequent comorbidity clusters, which were comprised of 14 latent features (LFs). Each LF was associated with a main disease (e.g., 98% of patients in cluster 2, characterized by LF2, had congestive heart failure [CHF]). The presence of certain LFs increased the probability of having another LF active. For example, LF2 (CHF) frequently appeared with LFs related to chronic kidney disease (CKD). Over time, the clusters associated with cardiovascular diseases, such as CHF, progressed rapidly. Moreover, the onset of certain diseases led to the appearance of further complications (e.g., CHF onset was associated with an increasing prevalence of CKD).

**Conclusions:** Our models identified established T2DM complications and previously unknown connections, thus, highlighting the potential for BNLMs t to characterize complex comorbidity patterns.

## Introduction

Once patients are diagnosed with Type 2 Diabetes Mellitus (T2DM) a constellation of chronic comorbidities might develop over time [1]. Common comorbidities are cardiovascular disease, diabetic retinopathy, peripheral neuropathy, and at later stages, chronic kidney disease (CKD) and musculoskeletal complications [2,3]. This implies that multimorbid T2DM patients have a high disease burden and are likely to experience a high degree of polypharmacy [4]. Understanding the development of comorbidities and identifying common trajectories may aid in the development of more personalized management strategies. However, the evolution of chronic comorbidities in patients with T2DM is poorly understood.

With the growing availability of large electronic healthcare records and advances in machine learning, different statistical models have been used to find clusters of T2DM patients with similar disease or comorbidity progression. For instance, Aguado and colleagues used network analysis [5] to identify the common subsequent comorbidity development following T2DM diagnosis while Khan et al. utilized network analysis to predict the progression of diabetes [6]. A study by Ahlqvist et al. [7], which was later replicated using clinical data by Dennis et al. [8], identified five different subgroups of T2DM glycaemic progression by using *k*-means hierarchical clustering, based on six variables. Importantly, all these studies found that the clusters were associated with diabetic complications such as kidney disease or retinopathy. However, no study to date has aimed at examined changes in comorbidity clusters over time following the start of T2DM.

Modelling comorbidity progression can help clinicians understand and prevent both poor health trajectories and potentially harmful polypharmacy. Previous studies have used latent class analysis (LCA) in healthcare data to broadly model multimorbidity trajectories of chronic diseases [9–11]. However, LCA models pose some limitations such as they assume that the number of features is known and that the features follow a Gaussian distribution. Hence, we propose that adopting a Bayesian nonparametric model might help overcome these limitations as they allow data to be modelled in an unspecified number of latent features [12,13]. Only few epidemiological studies have used this approach, for instance, in understanding comorbidities in patients with psychiatric disorders [14] or in suicide attempts [15]. However, Bayesian nonparametric models have never been used in electronic health records to understand T2DM disease progression.

Therefore, in this study we aimed to identify and describe the progression of common chronic comorbidities after T2DM onset using a Bayesian nonparametric model in a primary care electronic health records database.

## Methods

### Data Source

The IQVIA Medical Research Database UK (IMRD-UK) incorporates data supplied from The Health Improvement Network (THIN), which is a Cegedim database of anonymized electronic health records, generated from the daily record of General Practitioners (GPs). It includes data of more than 18 million patients from over 800 GP practices in the UK, and about 6% of the UK population. The database contains detailed information about patient characteristics (i.e., year of birth, sex, practice registration date, practice de-registration date, ethnicity), medical conditions (i.e., diagnoses with dates, referrals to hospitals, symptoms), medications (i.e., drug name, formulation, date, strengths, quantity, dosing instructions), in practice immunizations, laboratory tests, and results, and other patient-level data (i.e., smoking status, height, weight, alcohol use, pregnancy, birth, death dates). While the database contains all data on prescribed medications, those purchased over-the-counter or prescriptions issued during secondary care (e.g., hospitals) do not appear in the database. All medications are mapped according to the international anatomical therapeutic codes (ATC) classification system and the British National Formulary (BNF) code dictionary. For medical conditions, all diagnoses are coded according to the Read clinical code system, a comprehensive coding language with over 100,000 codes and comparable to the international classification of diseases (ICD) system. The protocol for this project was approved by the THIN scientific research council (reference number: 20SR062).

### Study population

To identify patients with T2DM we included all adult patients (age 18+) with a first-ever prescription of a non-insulin antidiabetic drug (NIAD) prescribed between January 1st 2006 and December 31st 2019. The date of first NIAD prescription defined the index date (start of follow-up). In order to identify new users, patients were required to have a minimum of one year of valid data collection prior to the first-ever prescription of a NIAD. Patients with a history of polycystic ovarian syndrome (PCOS), gestational diabetes, or insulin prescription prior to the index date were excluded since these conditions are treated with NIAD although not being necessarily T2DM patients.

### Chronic disease conditions

Chronic diseases were identified as conditions that last longer than one year and require medical attention [16]. We selected 58 distinct chronic comorbidities using Read Codes **Supplementary Table S1**, the clinical terminology used in General Practice in the UK in which each Read Code represents a term or short phrase which describes a health-related concept [17]. Read Codes were simplified to the third level i.e., first three letters of the Read Code, in order to encompass all the possible and small deviations from the main diagnosis. For example, a “conductive hearing loss”, with Read Code F590500, can be collapsed to “conductive deafness”, F590.11, or further summarised to “hearing loss”, F59.00. The selected chronic conditions were based on conditions from the Quality Outcome Framework (QOF) and previous studies on comorbidities commonly associated with T2DM [11,18,19]. Given the large heterogeneity in the pathogenesis and pathophysiology of cancer we grouped all diagnoses of neoplasms under one single category (Read Codes starting with B). We identified existing comorbidities if a patient had ever had a recorded diagnosis on or prior to the index date. Finally, in order to avoid convergence problems of the models, we selected those chronic comorbidities that had a prevalence higher than 1.0% both for males and females.

We created a longitudinal patient-disease binary matrix in discrete diabetes years (i.e., years elapsed between chronic disease onset and index date). Therefore, every row corresponded to a specific patient in a given year, and the columns corresponded to the comorbidities that the patient had developed in that time point. For model fitting, we selected the last observed period for each patient, thus, we ended up with a single row per patient which encoded the chronic comorbidities that the patient had developed.

### Statistical methods

Prior to model development, we summarized main patient characteristics at index date, stratified by sex. Latent feature models assume that there is an unknown low-dimensional representation of patients-disease [20]. Traditional methods are matrix factorization or latent Dirichlet allocation (LDA) [21]. However, these approaches require the number of latent features to retrieve be specified, and they are assumed to follow a specific distribution e.g., Gaussian distribution. An elegant solution to these issues is achieved by using Bayesian nonparametric models, such as a General Latent Feature Model (GLFM), by posing an Indian Buffet Process (IBP) as nonparametric prior over binary observation matrices [22]. This generated a binary matrix where columns represent a potentially unlimited number of features, while rows, representing patients, are finite. Therefore, GLFMs conduct latent feature analysis without pre-specifying the number of latent features. Each data point 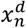 can be explained by a *K*-length binary vector **z**_*n*_ = [z_*n*1_, …, z_*nK*_] whose elements indicate whether a latent feature is active or not for the *n*^*th*^ object, and a real-valued weighting vector 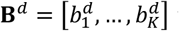 whose elements 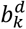 weight the influence of each latent feature in the *d*^*th*^ attribute of **X**. Therefore, the likelihood can be described as:

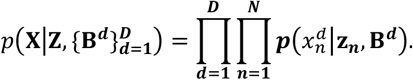

The binary latent feature vectors **z**_*n*_ are gathered in a *N* × *K* matrix **Z** which follows an IBP prior with *α* as a concentration parameter, i.e., **Z** ∼ *IBP*(*α*), where *α* controls the a priori activation probability of new features. Therefore, larger values will result in a higher number of expected latent features as well as a larger number of active features per row. For further details see [23]. Moreover, we forced the first latent feature to be always active, acting as a bias term i.e., all patients who do not have comorbidities or just one random, would only have the first latent feature, making this group to act as a baseline cluster.

On the **B**^*d*^ matrix we place a Gaussian prior, 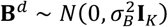. In order to overcome the problems of not having a Gaussian-distributed observation matrix, we transform each data point 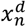 into an auxiliary Gaussian variable 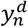, also called *pseudo-observation*, by applying a transformation function *f*_*d*_(·). The *pseudo-observation* is defined as

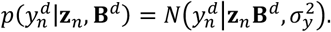

In the case of a binary observation matrix **X** each observation 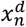 can only take two values 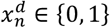. Hence, we can map the real values to the positive real numbers by applying the following transformation

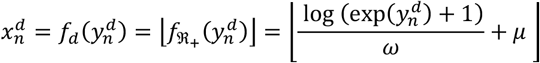

where *ω* and *μ* are scale and location hyper-parameters. Hence, the likelihood is defined as

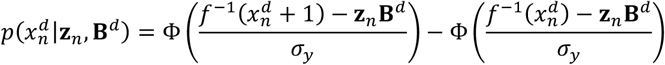

where 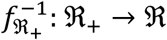 is the inverse function of the transformation 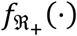.

### Inference

Given that the posterior distribution of **B**^*d*^ is intractable, we rely on a Markov Chain Monte Carlo (MCMC) approach, i.e., Gibbs sampling [23], to obtain posterior samples from Z and B. In order to speed up the sampling process, those patients who did not have any comorbidity were not sampled, and were assigned only the bias term. The sampling procedure can be summarized as follows:

Firstly, we sample **Z**

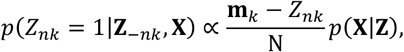

then we sample **B**^***d***^

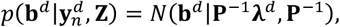

where 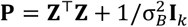 and **λ**^*d*^ = **Z**^⊤^**y**^*d*^. Finally, we sample *Y*^*d*^ given **X, Z, B**^d^,

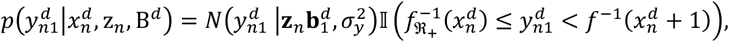

where we sample 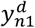 from a Gaussian left-truncated by 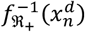 and right-truncated by 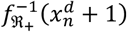. This inference procedure is repeated as many times as iterations set We set the Gibbs sampler to run for 1,000 iterations, the variance of the Gaussian prior to the weighing vectors **B**^**d**^ to 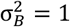, and the concentration parameter for the IBP to α = 1. In order to speed up the computations we did not sample those rows of **Z** corresponding to patients who did not have any disease.

### Predictions

In order to analyse the evolution of comorbidities over time we estimated the latent features that were active in each period per patient. To do so we retrieve all the unique combinations of latent features **z**_*i*_ from **Z** and compute the likelihood of each **z**_*i*_ to each observation *x*_*n*_, as previously shown,

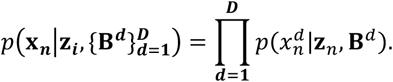

### Description of clusters

We described each cluster **z**_*i*_ and tabulated the count and proportion of patients with a specific disease within that cluster, the proportion of people with that specific disease on the overall population, and the Observed-Expected (O/E) ratio. The O/E ratio is defined as the ratio between proportion of patients with a given disease in a cluster divided the proportion of patients with that disease overall, and it gives a magnitude of how a specific comorbidity is over- or underrepresented in a given cluster. Moreover, we reported the proportion of females within that cluster and in the population overall, and computed the corresponding O/E ratio, the proportion of females in a cluster divided the proportion of females overall, to detect if there were female-dominated clusters.

We reported the empirical probabilities of possessing at least one latent feature or a single feature. Additionally, we computed the empirical and the product probability of possessing at least two latent features in order to identify if two given latent features were independent. For instance, once a latent feature is active the probability of having another given latent feature is higher. Finally, we also computed the probability of possessing at least latent features *k*_1_ and *k*_2_ given that *k*_1_ is active i.e.,

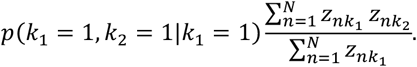

The Bayesian nonparametric model was implemented in C++, all statistical analyses and summary statistics were done in R version 3.5.1 (R Project for Statistical Computing). Network visualization was done in Gephi [24].

### Network visualization

To visualize the progression between clusters **z**_*i*_ over time we performed a network visualization. The history of latent membership for patient *n* in time *t* is represented by **Z**_*nt*_. Nodes represent the different clusters **z**_*i*_, and the directed edges the direction of the transition between clusters **z**_*i*_ in different times *t*. The size of the nodes, the weight, is proportional to the number of times that patients were in that specific node. In order to improve the visualization of the nodes we took the log of the node weight and rescale the weights between 0 and 1 as follows:

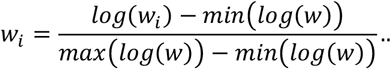

## Results

Following exclusions, a total of 175,383 eligible T2DM patients were identified, **Figure 1. Table 1** provides the demographic characteristics of the patients at index date, stratified by sex. There were 97,148 males and 78,235 females with an average age of 60.6 years old, both for males and females. The five most prevalent comorbidities at index date were high blood pressure (38.1), cancer (25.5%), osteoarthritis (19.8%), and anxiety and depression (17.2).

**Figure 1.**
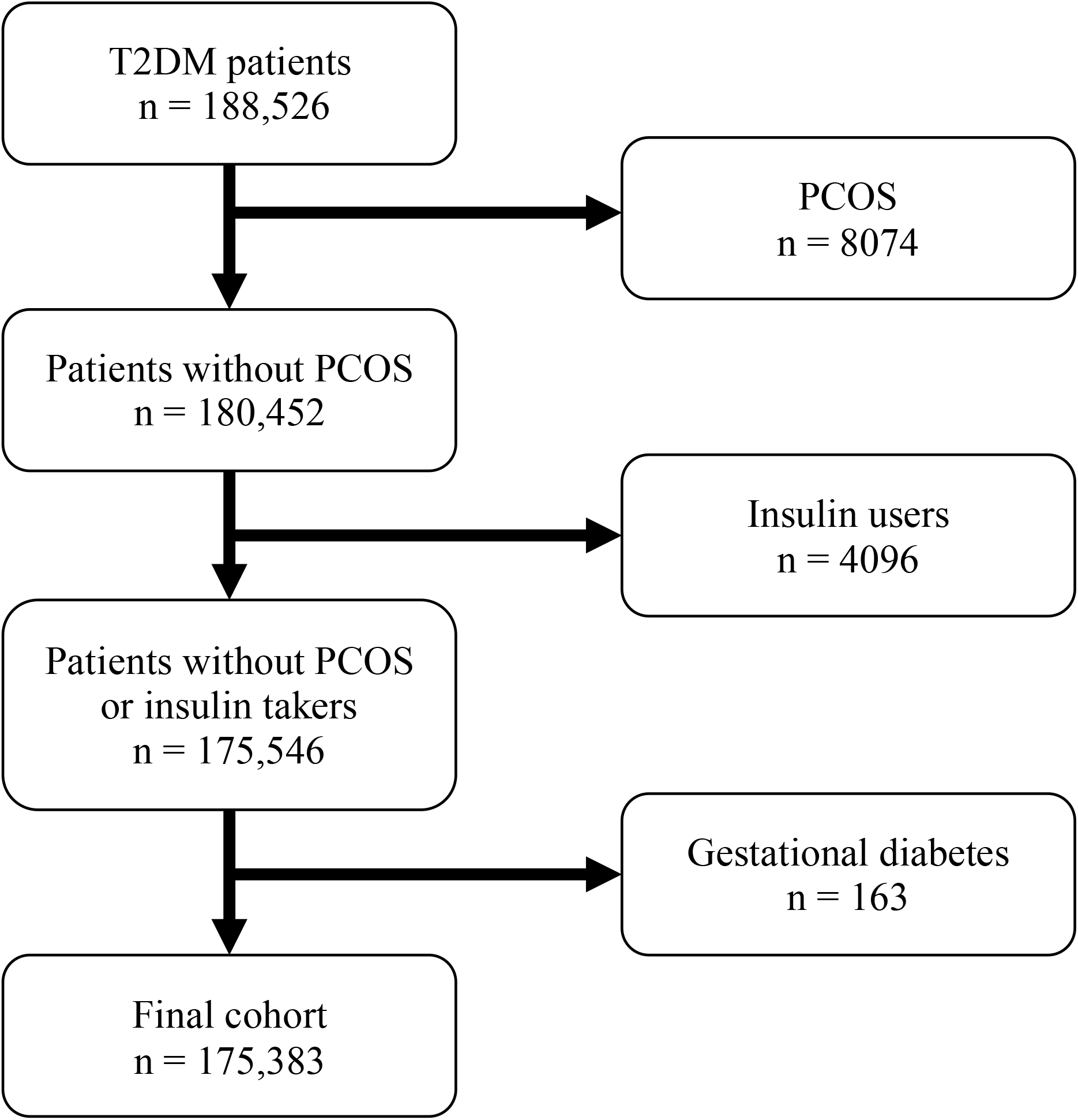
Flowchart of included patients. Abbreviations: T2DM, Type 2 diabetes mellitus; PCOS, polycystic ovarian syndrome;

**Table 1.** Demographic characteristics of 175,383 T2DM patients at index date (first NIAD prescription).

|  | **Overall**<br>(N=175383) | **Male**<br>(N=97148) | **Female**<br>(N=78235) | **SMD** |
| --- | --- | --- | --- | --- |
| Index Age (mean (SD)) | 60.6 (14.1) | 60.7 (12.8) | 60.4 (15.6) | 0.03 |
| Smoking (%) |  |  |  | 0.33 |
| Current | 60206 (34.5) | 39227 (40.6) | 20979 (26.9) |  |
| Never | 83719 (48.0) | 39715 (41.1) | 44004 (56.5) |  |
| Previous | 30641 (17.6) | 17729 (18.3) | 12912 (16.6) |  |
| Unkwown | <6 | <6 | <6 |  |
| Alcohol consumption (%) |  |  |  | 0.40 |
| Current | 7368 (4.5) | 4342 (4.8) | 3026 (4.2) |  |
| Never | 38687 (23.7) | 14679 (16.2) | 24008 (33.2) |  |
| Previous | 116823 (71.7) | 71470 (79.0) | 45353 (62.6) |  |
| Unkwown | 29 (0.0) | 16 (0.0) | 13 (0.0) |  |
| BMI (mean (SD)) | 32.54 (6.96) | 31.84 (6.19) | 33.39 (7.72) | 0.22 |
| Comorbidities (ever-before) |  |  |  |  |
| Cancer | 44670 (25.5) | 21275 (21.9) | 23395 (29.9) | 0.18 |
| Hypothyroidism | 13940 (7.9) | 3234 (3.3) | 10706 (13.7) | 0.38 |
| Pure hypercholesterolaemia | 28287 (16.1) | 16118 (16.6) | 12169 (15.6) | 0.03 |
| Obesity | 20051 (11.4) | 8760 (9.0) | 11291 (14.4) | 0.17 |
| Anxiety & other | 30167 (17.2) | 12273 (12.6) | 17894 (22.9) | 0.27 |
| Neuropathy | 1278 (0.7) | 769 (0.8) | 509 (0.7) | 0.02 |
| Primary open-angle glaucoma | 6072 (3.5) | 3252 (3.3) | 2820 (3.6) | 0.01 |
| Senile cataract | 8208 (4.7) | 3775 (3.9) | 4433 (5.7) | 0.08 |
| Deafness | 14323 (8.2) | 8487 (8.7) | 5836 (7.5) | 0.05 |
| High blood pressure | 66828 (38.1) | 36341 (37.4) | 30487 (39.0) | 0.03 |
| Angina pectoris | 11512 (6.6) | 7400 (7.6) | 4112 (5.3) | 0.10 |
| Atrial fibrillation | 12342 (7.0) | 7161 (7.4) | 5181 (6.6) | 0.03 |
| Congestive heart failure | 5016 (2.9) | 3135 (3.2) | 1881 (2.4) | 0.05 |
| Intermittent claudication | 5486 (3.1) | 3415 (3.5) | 2071 (2.6) | 0.05 |
| Chronic bronchitis | 2848 (1.6) | 1446 (1.5) | 1402 (1.8) | 0.02 |
| Irritable bowel syndrome | 17194 (9.8) | 6496 (6.7) | 10698 (13.7) | 0.23 |
| Chronic liver disease | 4345 (2.5) | 2407 (2.5) | 1938 (2.5) | 0.00 |
| Chronic kidney disease | 1070 (0.6) | 608 (0.6) | 462 (0.6) | 0.00 |
| Psoriasis or eczema | 10297 (5.9) | 5594 (5.8) | 4703 (6.0) | 0.01 |
| Osteoarthritis | 34683 (19.8) | 15915 (16.4) | 18768 (24.0) | 0.19 |
| Arthropathy | 9991 (5.7) | 4298 (4.4) | 5693 (7.3) | 0.12 |
| Cervical spondylosis | 13025 (7.4) | 6392 (6.6) | 6633 (8.5) | 0.07 |
| Osteoporosis | 7171 (4.1) | 2058 (2.1) | 5113 (6.5) | 0.22 |

Abbreviations: SD, standard deviation; SMD, Standardized mean difference. All identified comorbidities were identified via read codes, as recorded anytime on or before the index date, defined as the first prescribed non-insulin antidiabetic drug (NIAD).

From the initial list of 58 chronic conditions, we selected a total of 23 conditions that had a prevalence higher than 1.0% in order to avoid numerical and convergence problems of the Bayesian nonparametric model. The selected chronic comorbidities are shown in **Supplementary Table S1**.

We found 14 different latent features, of which the first one, the bias term, was active for every patient. The 14 latent features resulted in 385 clusters, with each cluster corresponding to a unique combination of the latent features. **Table 2** provides an overview of the 20 most common clusters and the top three most prevalent conditions associated to each. With the exception of cluster 1, which includes the bias term (i.e., latent feature 1), most of the clusters were represented by one highly prevalent chronic disease with other additional diseases having elevated O/E ratios. Therefore, for any given main disease, corresponding to a specific latent feature, some secondary comorbidities with a higher probability of onset were associated to it. For example, the second cluster, which had latent feature 2 active, was strongly associated with congestive heart failure (CHF), **Table 2**. Overall, 98% of the patients in the second cluster had CHF and the O/E ratio was 43.4. Additionally, once a patient developed CHF, the probability of concomitantly having atrial fibrillation and senile cataract was increased 7.0- and 2.3-fold, respectively, as seen in the corresponding O/E ratios. The third cluster was mainly composed of patients with hypothyroidism, while the fourth and fifth were characterized by patients with osteoporosis and obesity, respectively.

**Table 2.** Description of the three most prevalent conditions for the first 20 clusters.

| Cluster | Latent features | Recorded comorbidities | Count | N cluster | Count prop | Total disease | Total dis. Prop | O/E ratio |
| --- | --- | --- | --- | --- | --- | --- | --- | --- |
| 1 | LF1 | High blood pressure | 8396 | 147816 | 5.7% | 10660 | 6.1% | 0.9 |
| 1 | LF1 | Pure hypercholesterolaemia | 4570 | 147816 | 3.1% | 5938 | 3.4% | 0.9 |
| 1 | LF1 | Chronic liver disease | 3437 | 147816 | 2.3% | 4505 | 2.6% | 0.9 |
| 2 | LF1 LF2 | Congestive heart failure | 2711 | 2766 | 98.0% | 3962 | 2.3% | 43.4 |
| 2 | LF1 LF2 | Atrial fibrillation | 776 | 2766 | 28.1% | 7000 | 4.0% | 7.0 |
| 2 | LF1 LF2 | Senile cataract | 281 | 2766 | 10.2% | 7709 | 4.4% | 2.3 |
| 3 | LF1 LF3 | Hypothyroidism | 2535 | 2572 | 98.6% | 3260 | 1.9% | 53.0 |
| 3 | LF1 LF3 | Irritable bowel syndrome | 118 | 2572 | 4.6% | 5019 | 2.9% | 1.6 |
| 3 | LF1 LF3 | Anxiety & other | 92 | 2572 | 3.6% | 4115 | 2.3% | 1.5 |
| 4 | LF1 LF4 | Osteoporosis | 2323 | 2350 | 98.9% | 3233 | 1.8% | 53.6 |
| 4 | LF1 LF4 | Senile cataract | 269 | 2350 | 11.4% | 7709 | 4.4% | 2.6 |
| 4 | LF1 LF4 | Irritable bowel syndrome | 148 | 2350 | 6.3% | 5019 | 2.9% | 2.2 |
| 5 | LF1 LF5 | Obesity | 2235 | 2252 | 99.2% | 2879 | 1.6% | 60.5 |
| 5 | LF1 LF5 | Pure hypercholesterolaemia | 185 | 2252 | 8.2% | 5938 | 3.4% | 2.4 |
| 5 | LF1 LF5 | Chronic liver disease | 121 | 2252 | 5.4% | 4505 | 2.6% | 2.1 |
| 6 | LF1 LF6 | Intermittent claudication | 1855 | 1871 | 99.1% | 2653 | 1.5% | 65.5 |
| 6 | LF1 LF6 | Atrial fibrillation | 153 | 1871 | 8.2% | 7000 | 4.0% | 2.1 |
| 6 | LF1 LF6 | Senile cataract | 153 | 1871 | 8.2% | 7709 | 4.4% | 1.9 |
| 7 | LF1 LF7 | Primary open-angle glaucoma | 1789 | 1795 | 99.7% | 2330 | 1.3% | 75.0 |
| 7 | LF1 LF7 | Senile cataract | 289 | 1795 | 16.1% | 7709 | 4.4% | 3.7 |
| 7 | LF1 LF7 | Osteoarthritis | 143 | 1795 | 8.0% | 8954 | 5.1% | 1.6 |
| 8 | LF1 LF9 | Arthropathy | 1531 | 1532 | 99.9% | 2168 | 1.2% | 80.8 |
| 8 | LF1 LF9 | Osteoarthritis | 290 | 1532 | 18.9% | 8954 | 5.1% | 3.7 |
| 8 | LF1 LF9 | Anxiety & other | 70 | 1532 | 4.6% | 4115 | 2.3% | 2.0 |
| 9 | LF1 LF8 | Chronic bronchitis | 1494 | 1496 | 99.9% | 2260 | 1.3% | 77.5 |
| 9 | LF1 LF8 | Deafness | 95 | 1496 | 6.4% | 5261 | 3.0% | 2.1 |
| 9 | LF1 LF8 | Senile cataract | 134 | 1496 | 9.0% | 7709 | 4.4% | 2.0 |
| 10 | LF1 LF10 | Psoriasis or eczema | 1477 | 1481 | 99.7% | 1975 | 1.1% | 88.6 |
| 10 | LF1 LF10 | Osteoarthritis | 144 | 1481 | 9.7% | 8954 | 5.1% | 1.9 |
| 10 | LF1 LF10 | Chronic liver disease | 71 | 1481 | 4.8% | 4505 | 2.6% | 1.9 |

**Table 2. (continued)** Description of the three most prevalent conditions for the first 20 clusters.
| Cluster | Latent features | Recorded comorbidities | Count | N cluster | Count prop | Total disease | Total dis. Prop | O/E ratio |
| --- | --- | --- | --- | --- | --- | --- | --- | --- |
| 11 | LF1 LF12 | Cervical spondylosis | 1369 | 1371 | 99.9% | 1937 | 1.1% | 90.4 |
| 11 | LF1 LF12 | Osteoarthritis | 204 | 1371 | 14.9% | 8954 | 5.1% | 2.9 |
| 11 | LF1 LF12 | Irritable bowel syndrome | 97 | 1371 | 7.1% | 5019 | 2.9% | 2.5 |
| 12 | LF1 LF11 | Neuropathy | 1350 | 1350 | 100.0% | 1964 | 1.1% | 89.3 |
| 12 | LF1 LF11 | Chronic liver disease | 65 | 1350 | 4.8% | 4505 | 2.6% | 1.9 |
| 12 | LF1 LF11 | Irritable bowel syndrome | 70 | 1350 | 5.2% | 5019 | 2.9% | 1.8 |
| 13 | LF1 LF14 | Angina pectoris | 1253 | 1255 | 99.8% | 1773 | 1.0% | 98.8 |
| 13 | LF1 LF14 | Deafness | 80 | 1255 | 6.4% | 5261 | 3.0% | 2.1 |
| 13 | LF1 LF14 | Atrial fibrillation | 105 | 1255 | 8.4% | 7000 | 4.0% | 2.1 |
| 14 | LF1 LF13 | Chronic kidney disease | 1227 | 1228 | 99.9% | 1869 | 1.1% | 93.8 |
| 14 | LF1 LF13 | Atrial fibrillation | 109 | 1228 | 8.9% | 7000 | 4.0% | 2.2 |
| 14 | LF1 LF13 | Deafness | 81 | 1228 | 6.6% | 5261 | 3.0% | 2.2 |
| 15 | LF1 LF13 LF2 | Chronic kidney disease | 135 | 135 | 100.0% | 1869 | 1.1% | 93.8 |
| 15 | LF1 LF13 LF2 | Congestive heart failure | 135 | 135 | 100.0% | 3962 | 2.3% | 44.3 |
| 15 | LF1 LF13 LF2 | Atrial fibrillation | 45 | 135 | 33.3% | 7000 | 4.0% | 8.4 |
| 16 | LF1 LF8 LF2 | Chronic bronchitis | 126 | 126 | 100.0% | 2260 | 1.3% | 77.6 |
| 16 | LF1 LF8 LF2 | Congestive heart failure | 125 | 126 | 99.2% | 3962 | 2.3% | 43.9 |
| 16 | LF1 LF8 LF2 | Atrial fibrillation | 49 | 126 | 38.9% | 7000 | 4.0% | 9.7 |
| 17 | LF1 LF4 LF2 | Osteoporosis | 117 | 117 | 100.0% | 3233 | 1.8% | 54.3 |
| 17 | LF1 LF4 LF2 | Congestive heart failure | 117 | 117 | 100.0% | 3962 | 2.3% | 44.3 |
| 17 | LF1 LF4 LF2 | Atrial fibrillation | 29 | 117 | 24.8% | 7000 | 4.0% | 6.2 |
| 18 | LF1 LF3 LF2 | Hypothyroidism | 103 | 103 | 100.0% | 3260 | 1.9% | 53.8 |
| 18 | LF1 LF3 LF2 | Congestive heart failure | 103 | 103 | 100.0% | 3962 | 2.3% | 44.3 |
| 18 | LF1 LF3 LF2 | Atrial fibrillation | 35 | 103 | 34.0% | 7000 | 4.0% | 8.5 |
| 19 | LF1 LF6 LF2 | Intermittent claudication | 101 | 101 | 100.0% | 2653 | 1.5% | 66.1 |
| 19 | LF1 LF6 LF2 | Congestive heart failure | 101 | 101 | 100.0% | 3962 | 2.3% | 44.3 |
| 19 | LF1 LF6 LF2 | Atrial fibrillation | 32 | 101 | 31.7% | 7000 | 4.0% | 7.9 |
| 20 | LF1 LF14 LF2 | Angina pectoris | 96 | 96 | 100.0% | 1773 | 1.0% | 98.9 |
| 20 | LF1 LF14 LF2 | Congestive heart failure | 96 | 96 | 100.0% | 3962 | 2.3% | 44.3 |
| 20 | LF1 LF14 LF2 | Atrial fibrillation | 25 | 96 | 26.0% | 7000 | 4.0% | 6.5 |

Abbreviations: N Cluster, number of patients within that cluster; Count prop, proportion of patients who had the disease within that cluster; Total disease, total number of patients with the disease; Total dis. Prop, proportion of patients who have the disease, overall; O/E Ratio, Observed-to-Expected ratio;
All identified comorbidities were identified via read codes, as recorded anytime on or before the index date, defined as the first prescribed non-insulin antidiabetic drug (NIAD)

From cluster 15 on, we observed that the clusters resulted from the combination of two or more latent features, **Table 2**. Hence, patients had two distinct main diseases along with other secondary comorbidities. For instance, cluster 15 was composed of patients who all (100%) had chronic kidney disease (CKD) and CHF, while one-third (33.3%) had atrial fibrillation. All three top conditions also had elevated O/E ratios of 93.8, 44.3, and 8.4, respectively. In cluster 16, all (100%) patients had chronic bronchitis, while 99.2% also had CHF, and 38.9% had atrial fibrillation. Again, elevated O/E ratios were identified for the three top conditions. The complete list of comorbidities identified per cluster are provided in **Supplementary Table S2**.

Sex differences between clusters were also identified as shown in **Supplementary Figure S1**. For example, cluster 2 and 6 (cardiovascular disease clusters) were more heavily dominated by males, as evidenced by the lower proportion of females within the clusters (34.8% and 33.1% female, respectively). Similarly, the O/E ratios for the gender distribution was below 1.0 in both clusters (e.g., 0.7 and 0.78, for cluster 2 and 6, respectively). Conversely, other clusters were female dominated. For example, clusters 3 and 4 (hypothyroidism and osteoporosis) consisted of 57.8% and 71.3% females, respectively, and the sex O/E ratios were 1.3 and 1.6, respectively), **Supplementary Figure S1**.

In **Table 3**, we present the probability of presenting at least one of the latent features active, either in combination with other latent features or as a single feature. We found that 84.3% of the individuals had only the bias term, latent feature 1, active. Moreover, certain comorbidities were more likely to appear than others. For instance, latent feature 2, corresponding to CHF, was the feature with highest probability of being active, either in combination with other features (2.3%), or as a single feature (1.6%). The least likely features to be active, either in combination with others or as a single feature, were latent feature 13 and 14, associated with chronic kidney disease and angina pectoris respectively, as shown in **Table 3**. Hence, having CHF along with its comorbidities was more likely than having CKD with other comorbidities.

**Table 3.** Probabilities (%) of possessing at least one latent feature, or a single feature.

| Latent Feature | Total | Single Feature | Dominant Feature |
| --- | --- | --- | --- |
| 1 | 100% | 84.28% | High blood pressure |
| 2 | 2.30% | 1.58% | Congestive heart failure |
| 3 | 1.89% | 1.47% | Hypothyroidism |
| 4 | 1.87% | 1.34% | Irritable bowel syndrome |
| 5 | 1.65% | 1.28% | Obesity |
| 6 | 1.53% | 1.07% | Intermittent claudication |
| 7 | 1.34% | 1.02% | Primary open-angle glaucoma |
| 8 | 1.29% | 0.85% | Arthropathy |
| 9 | 1.24% | 0.87% | Chronic bronchitis |
| 10 | 1.13% | 0.84% | Psoriasis or eczema |
| 11 | 1.12% | 0.77% | Cervical spondylosis |
| 12 | 1.11% | 0.78% | Neuropathy |
| 13 | 1.07% | 0.70% | Angina pectoris |
| 14 | 1.01% | 0.72% | Chronic kidney disease |

The probability of having at least two latent features active is presented in **Table 4**. We found that the empirical probability of two latent features was around twice as large as the product probabilities, indicating that an active latent feature was associated with an increased probability of having another latent feature active. For instance, the empirical probability of having latent feature 2 active, which is dominated by a high prevalence of CHF, and latent feature 4, associated with osteoporosis, was 0.11%, which was 2.5 times higher than the product probability of 0.04%.

**Table 4.** Probabilities of possessing at least two latent features. The elements below the diagonal correspond to the empirical probability, and the elements above the diagonal correspond to the product probability*.

| Latent Features | 1 | 2 | 3 | 4 | 5 | 6 | 7 | 8 | 9 | 10 | 11 | 12 | 13 | 14 |
| --- | --- | --- | --- | --- | --- | --- | --- | --- | --- | --- | --- | --- | --- | --- |
| 1 |  | 2.30% | 1.89% | 1.87% | 1.65% | 1.53% | 1.34% | 1.29% | 1.24% | 1.13% | 1.12% | 1.11% | 1.07% | 1.01% |
| 2 | 2.30% |  | 0.04% | 0.04% | 0.04% | 0.04% | 0.03% | 0.03% | 0.03% | 0.03% | 0.03% | 0.03% | 0.02% | 0.02% |
| 3 | 1.89% | 0.08% |  | 0.04% | 0.03% | 0.04% | 0.03% | 0.02% | 0.02% | 0.02% | 0.02% | 0.02% | 0.02% | 0.02% |
| 4 | 1.87% | 0.11% | 0.07% |  | 0.03% | 0.06% | 0.05% | 0.02% | 0.02% | 0.03% | 0.04% | 0.06% | 0.02% | 0.02% |
| 5 | 1.65% | 0.07% | 0.04% | 0.03% |  | 0.03% | 0.02% | 0.02% | 0.02% | 0.03% | 0.02% | 0.04% | 0.02% | 0.02% |
| 6 | 1.53% | 0.10% | 0.03% | 0.03% | 0.03% |  | 0.02% | 0.02% | 0.02% | 0.02% | 0.02% | 0.02% | 0.02% | 0.02% |
| 7 | 1.34% | 0.05% | 0.04% | 0.02% | 0.03% | 0.04% |  | 0.02% | 0.02% | 0.02% | 0.01% | 0.03% | 0.01% | 0.01% |
| 8 | 1.29% | 0.11% | 0.04% | 0.08% | 0.04% | 0.06% | 0.02% |  | 0.04% | 0.03% | 0.03% | 0.03% | 0.01% | 0.01% |
| 9 | 1.24% | 0.07% | 0.03% | 0.05% | 0.05% | 0.04% | 0.03% | 0.02% |  | 0.04% | 0.03% | 0.04% | 0.01% | 0.01% |
| 10 | 1.13% | 0.03% | 0.03% | 0.02% | 0.02% | 0.02% | 0.02% | 0.01% | 0.01% |  | 0.01% | 0.03% | 0.01% | 0.01% |
| 11 | 1.12% | 0.04% | 0.03% | 0.02% | 0.03% | 0.07% | 0.03% | 0.01% | 0.01% | 0.03% |  | 0.03% | 0.01% | 0.01% |
| 12 | 1.11% | 0.04% | 0.03% | 0.02% | 0.02% | 0.03% | 0.01% | 0.01% | 0.01% | 0.01% | 0.01% |  | 0.01% | 0.01% |
| 13 | 1.07% | 0.12% | 0.05% | 0.05% | 0.03% | 0.04% | 0.02% | 0.03% | 0.03% | 0.02% | 0.03% | 0.03% |  | 0.01% |
| 14 | 1.01% | 0.08% | 0.02% | 0.04% | 0.03% | 0.04% | 0.02% | 0.03% | 0.02% | 0.02% | 0.03% | 0.03% | 0.02% |  |

Empirical probabilities were directly extracted from the observed latent matrix.
\*Product probabilities were calculated from the individual latent feature probabilities, as reported in Table 3.

Additionally, we also saw that some diseases increased the probability of having concomitantly other diseases. Having a given feature active lead to an increased probability of having another one active, **Table 5**. For example, latent feature 2 appeared frequently with features 4, 6, 8, 13, and 14. Therefore, this would indicate that osteoporosis, intermittent claudication, arthropathy, angina pectoris, and CKD, were commonly associated with CHF in our T2DM cohort.

**Table 5.**
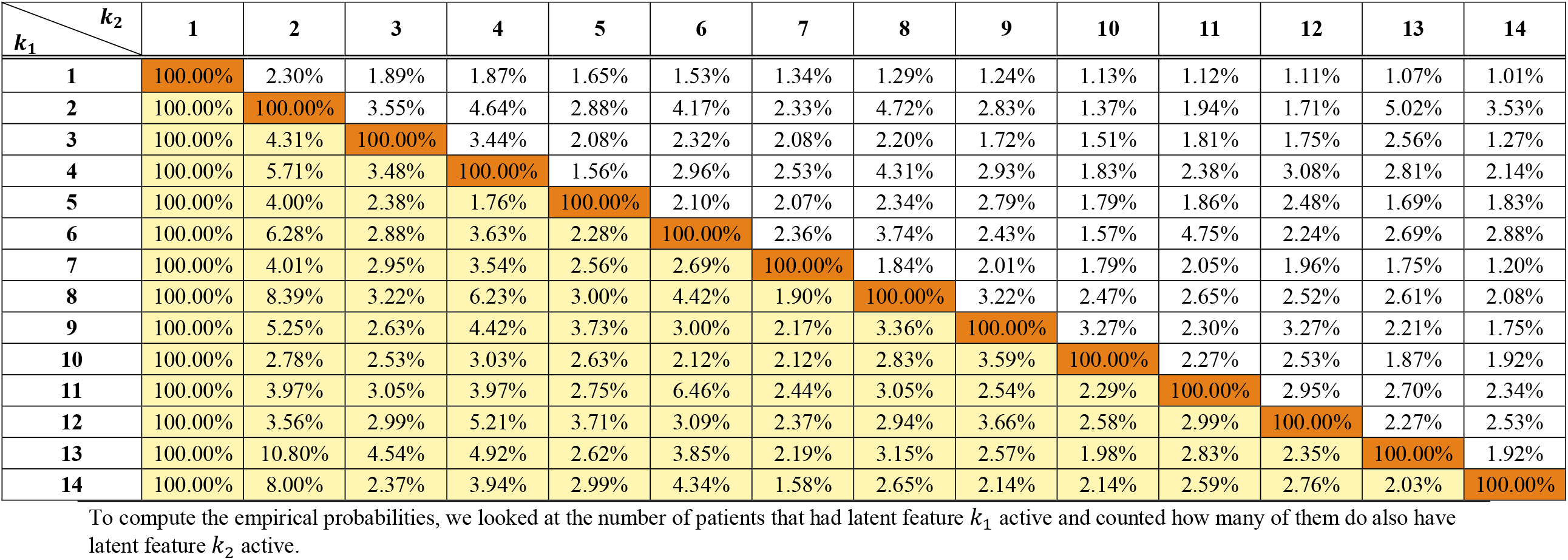
Empirical probabilities of possessing at least features *k*_1_ and *k*_2_ given that *k*_1_ is active.

| $k_1 \backslash k_2$ | 1 | 2 | 3 | 4 | 5 | 6 | 7 | 8 | 9 | 10 | 11 | 12 | 13 | 14 |
| --- | --- | --- | --- | --- | --- | --- | --- | --- | --- | --- | --- | --- | --- | --- |
| 1 | 100.00% | 2.30% | 1.89% | 1.87% | 1.65% | 1.53% | 1.34% | 1.29% | 1.24% | 1.13% | 1.12% | 1.11% | 1.07% | 1.01% |
| 2 | 100.00% | 100.00% | 3.55% | 4.64% | 2.88% | 4.17% | 2.33% | 4.72% | 2.83% | 1.37% | 1.94% | 1.71% | 5.02% | 3.53% |
| 3 | 100.00% | 4.31% | 100.00% | 3.44% | 2.08% | 2.32% | 2.08% | 2.20% | 1.72% | 1.51% | 1.81% | 1.75% | 2.56% | 1.27% |
| 4 | 100.00% | 5.71% | 3.48% | 100.00% | 1.56% | 2.96% | 2.53% | 4.31% | 2.93% | 1.83% | 2.38% | 3.08% | 2.81% | 2.14% |
| 5 | 100.00% | 4.00% | 2.38% | 1.76% | 100.00% | 2.10% | 2.07% | 2.34% | 2.79% | 1.79% | 1.86% | 2.48% | 1.69% | 1.83% |
| 6 | 100.00% | 6.28% | 2.88% | 3.63% | 2.28% | 100.00% | 2.36% | 3.74% | 2.43% | 1.57% | 4.75% | 2.24% | 2.69% | 2.88% |
| 7 | 100.00% | 4.01% | 2.95% | 3.54% | 2.56% | 2.69% | 100.00% | 1.84% | 2.01% | 1.79% | 2.05% | 1.96% | 1.75% | 1.20% |
| 8 | 100.00% | 8.39% | 3.22% | 6.23% | 3.00% | 4.42% | 1.90% | 100.00% | 3.22% | 2.47% | 2.65% | 2.52% | 2.61% | 2.08% |
| 9 | 100.00% | 5.25% | 2.63% | 4.42% | 3.73% | 3.00% | 2.17% | 3.36% | 100.00% | 3.27% | 2.30% | 3.27% | 2.21% | 1.75% |
| 10 | 100.00% | 2.78% | 2.53% | 3.03% | 2.63% | 2.12% | 2.12% | 2.83% | 3.59% | 100.00% | 2.27% | 2.53% | 1.87% | 1.92% |
| 11 | 100.00% | 3.97% | 3.05% | 3.97% | 2.75% | 6.46% | 2.44% | 3.05% | 2.54% | 2.29% | 100.00% | 2.95% | 2.70% | 2.34% |
| 12 | 100.00% | 3.56% | 2.99% | 5.21% | 3.71% | 3.09% | 2.37% | 2.94% | 3.66% | 2.58% | 2.99% | 100.00% | 2.27% | 2.53% |
| 13 | 100.00% | 10.80% | 4.54% | 4.92% | 2.62% | 3.85% | 2.19% | 3.15% | 2.57% | 1.98% | 2.83% | 2.35% | 100.00% | 1.92% |
| 14 | 100.00% | 8.00% | 2.37% | 3.94% | 2.99% | 4.34% | 1.58% | 2.65% | 2.14% | 2.14% | 2.59% | 2.76% | 2.03% | 100.00% |

To compute the empirical probabilities, we looked at the number of patients that had latent feature $k_1$ active and counted how many of them do also have latent feature $k_2$ active.

Complementarily, we compared the three main clusters associated with cardiovascular disease, **Supplementary Table S3**. We present the proportion of patients with each comorbidity overall and within the three clusters. Additionally, the O/E ratios by cluster are provided. Additionally, we compare the proportions for each comorbidity across clusters, using cluster 2 as the comparator. Cluster 2 was characterized by latent feature 2, CHF, and associated with higher prevalence of atrial fibrillation and senile cataract. However, when latent feature 13 was also active, cluster 15, a slight shift was observed. Here, 100% of the individuals had CHF and CKD. Moreover, most of the O/E ratios increased in this cluster, when compared to cluster 2; except for deafness, irritable bowel syndrome, anxiety, and chronic liver disease. Similarly, when latent features 2 and 8 were active in cluster 16, 100% of the patients had chronic bronchitis and again most of the O/E ratios were increased. Hence, the more active latent features active the sicker the patients, since they were characterised by two or more main disease e.g. CHF and CKD, and their respective secondary diseases.

In **Figure 2A**, we visualize the progression of the top 20 clusters by estimating the proportion of patients belonging to the individual clusters over time. As the proportion of people belonging to each cluster increased over time, we can identify that the probability of belonging to the baseline cluster steadily decreased over time, **Figure 2A**. Similarly, when looking at the 14 individual latent features in **Figure 2B**, we found that the probability of having a given latent feature active increased over time, except for the first latent feature (not shown) which was always active with a constant probability of 1. While the prevalence of latent feature 3 and latent feature 5, which corresponded to hypothyroidism and obesity, were highest at baseline, we observe that the proportion of patients with latent feature 2, which was associated with a high prevalence of CHF, increased at a higher rate compared to the other features. We also see an increase in the prevalence of latent feature 4, osteoporosis, over time **Figure 2B**.

**Figure 2A.**
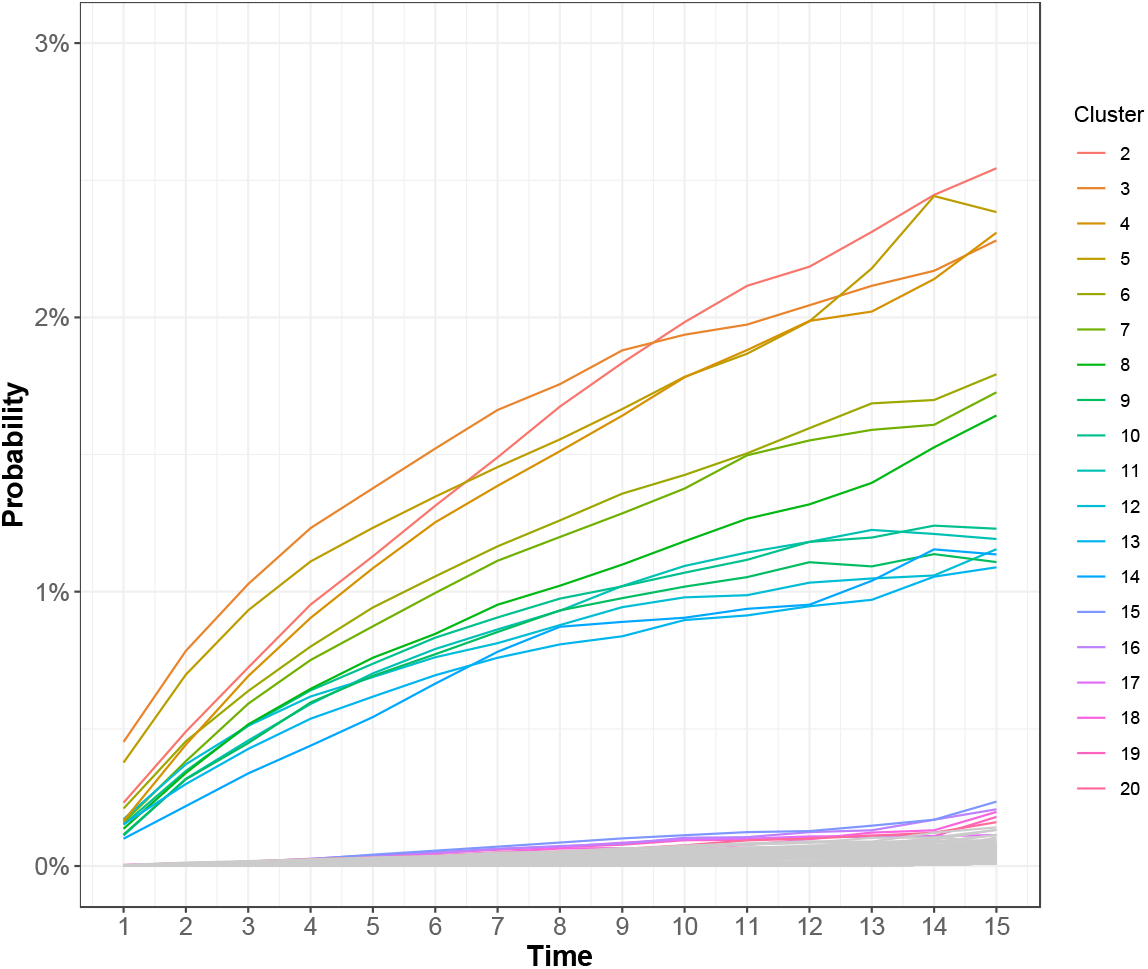
Probability of belonging to each cluster over time. More information on each cluster characteristics can be found on Table 2. Note that some clusters increased at a higher rate compared to others.

**Figure 2B.**
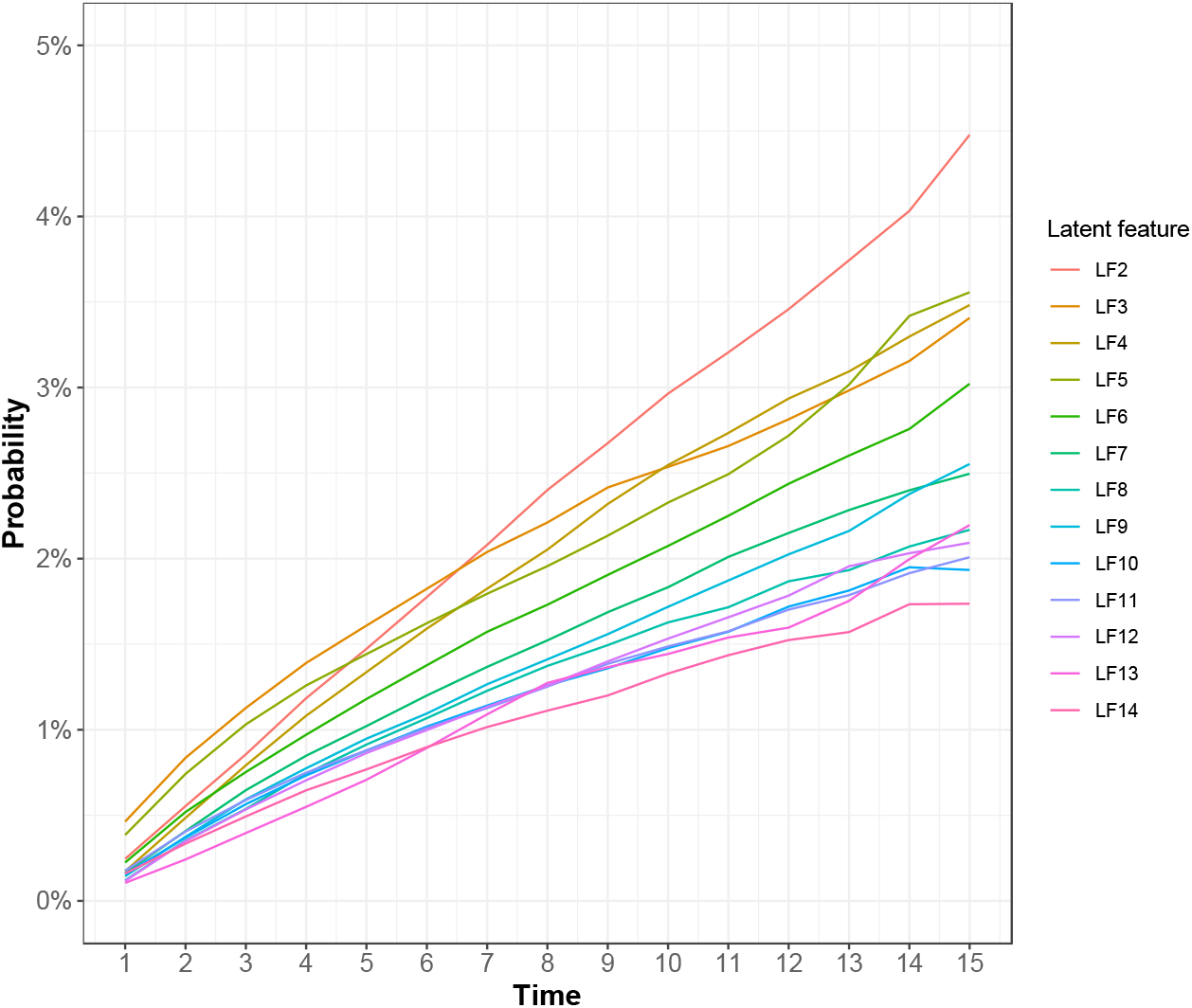
Evolution of active latent features over time. Latent feature 1 is not depicted as it is always active.

The network analysis that depicts the transition between the top 20 clusters over time is provided in **Supplementary Figure S2**. Overall patients tended to remain in the same cluster over time, however, transitions from the first (latent feature 1 active) cluster the other 14 (which are characterized by a single latent feature) were the most frequent. We further note that patients in clusters 5, 7, 8, 10-12 did not transition to other more complex clusters over time. However, a transition into cluster 2, was associated with further transitions into clusters 15-20, which were characterized by the presence of 2 active latent features.

## Discussion

This study confirmed the potential of using a large electronic healthcare database to identify clusters of chronic disease comorbidities among patients with newly treated T2DM. This is the first analysis that applied a Bayesian nonparametric model to real-world electronic medical records to identify distinct comorbidity clusters and disease progression patterns based on hidden latent features. In our case example, of patients with T2DM, we could identify 14 different latent features, in which each of them was strongly associated with a main disease. Importantly, we were able to identify comorbidity patterns consistent with the literature, pointing to the applicability of this approach in medical data. Thus, we found that Bayesian non-parametric models are a powerful tool to use in electronic health records to identify unique comorbidity clusters and health trajectories.

Understanding disease progression in T2DM patients is paramount in order to prevent disease onset, optimize treatment strategies, reduce polypharmacy, and in turn, increase safety and efficacy of therapeutic options. However, due to the complexity of comorbidity patterns, there is a lack of understanding of common patterns or trajectories. Previous studies have modelled T2DM progression in electronic health records using different approaches, including network modelling [6] naïve Bayes, support vector machines, random forests, and gradient boosted trees [25,26], or by using typical and atypical disease trajectory analysis [27]. Although these approaches can shed some light on the disease progression and comorbidities development, they might not be able to capture relationships between hidden or unknown risk factors. While the use of latent feature models can overcome important shortcomings of the aforementioned approaches, most models require to pre-specify the number of latent features to be retrieved. Consequently, they might not perform very well in the presence of binary matrices, and might lack interpretability because latent feature might extend over the real line [14,15].

The results of our study, identify that a Bayesian nonparametric model is a novel approach for studying chronic comorbidity progression. Bayesian nonparametric models overcome the limitations of traditional latent feature models as they can automatically infer the number of binary latent features from the data [23]. Using this approach, we found that the development of certain comorbidities can lead to a dramatic increase in the probabilities of developing other conditions over time. For example, in our analysis, once a patient with T2DM developed CHF, their probability of being diagnosed with atrial fibrillation increases as seen in cluster 2. We also found that patients with hypothyroidism had an elevated likelihood of being diagnosed with irritable bowel syndrome as well as anxiety and neurotic disorders increase [28,29]. While previous literature has found individual associations between hypothyroidism, irritable bowel syndrome, and anxiety [30,31], the link between these as a common cluster, particularly among patients with T2DM, has not been previously identified. Therefore, our models were able to identify hidden (or previously unknown) connections between the diseases that form each cluster.

While our models were able to identify unique comorbidity clusters, the predicted posterior probabilities were consistent with the known progression of T2DM. For instance, we found that all latent features, especially latent feature 2, which was associated with cardiovascular events, steadily increased over time. Conversely, the posterior probability for the baseline cluster, only latent feature 1 active, decreased over time. These results are in line with previous literature, for instance Khan et al. found that cardiovascular conditions such as cardiac arrythmias or hypertension were the most prevalent diseases appearing after T2DM onset [6] or Oh and colleagues who identified hyperlipidaemia and hypertension as frequent comorbidities after T2DM diagnosis [27]. Hence, after T2DM onset the probability of developing certain comorbidities increases over the course of the disease.

### Limitations

Although our study was population-based and applied Bayesian nonparametric models, which overcome many of the limitations found in previous work, there are remaining limitations that must be considered when interpreting the results of this study. Firstly, we only looked at a specific subset of 23 different chronic comorbidities, thus, we might have missed some patterns in the data. Moreover, we did not include acute outcomes in our list of comorbidities. We acknowledge that chronic diseases can increase the risk of experiencing an acute event, similarly, acute events can also trigger or accelerate the onset of new chronic conditions. Therefore, future research could assess if similar trajectories are found when incorporating acute events, or the impact of the chronic disease clusters on the onset of new acute outcomes. In addition, since comorbidities were binary encoded, and remained active after the first diagnosis, we might have missed different severity levels that a chronic disease might have had. Moreover, we did not include pharmacological treatments, which can have an impact in the onset/delay of new comorbidities or alter current disease status.

Cancer is a very complex and heterogenous disease that requires thorough medical attention. In our analysis we grouped all cancer diagnoses under one single code for the sake of interpretability. Nonetheless, there might have missed specific links between different cancer types and comorbidity clusters, particularly those that are more commonly associated with T2DM (e.g., pancreatic or gastric cancer). Therefore, future studies may consider using Bayesian nonparametric models to investigate comorbidity clusters associated with specific cancer diagnoses to generate new hypotheses in diabetes patients.

## Conclusions

In this population-based study of patient with T2DM, we were able to confirm the potential of using a Bayesian nonparametric model to identify distinct patient clusters. Our models found results that are consistent with literature (e.g., growing prevalence of cardiovascular disease), thereby providing confidence in the utility. In contrast to previous studies based on latent feature analysis, we were able to uncover previously unknown, or hidden, factors. Based on these results, Bayesian nonparametric models may be useful to develop our understanding of complex comorbidity patterns and disease progression in chronic diseases. Deeper understanding of T2DM progression and multimorbidity can foster new hypotheses for further epidemiological studies and be used in clinical guidance of the patients.

## Data Availability

The IQVIA Medical Research Database UK (IMRD-UK) incorporates data supplied from The Health Improvement Network (THIN), which is a Cegedim database of anonymized electronic health records, generated from the daily record of General Practitioners (GPs).

## Author Contributions

Study Conception: AMB, FPC; data acquisition: AMB; data analysis: AMDlT, FPC; data integrity and validity: AMDlT, AMB; data interpretation: AMDlT, FPC, SW, AMB; manuscript preparation: AMDlT, AMB; critical revisions: AMDlT, FPC, SW, AMB.

## Acknowledgments

The authors would like to acknowledge the following individuals for their assistance in the project. Dr. Guillaume Obozinski, Dr. Victor Cohen, Dr. Ekaterina Krymova, Dr. Esra Suel, Dr. Izabela Moise, and Dr. Anna Susmelj for the input and methodological discussions. Dr. Sofiane Sarni and Fotis Georgatos for software assistance. Dr. Melanie Fernandez Pradier for her invaluable help in adapting and implementing the models.

