## Supplementary Material for "Comorbidity clusters associated with newly treated Type 2 diabetes mellitus: a Bayesian nonparametric analysis"

### Title

### Author Names and Affiliations

Adrian Martinez-De la Torre[1], MSc

Fernando Perez-Cruz[2,3], PhD

Stefan Weiler[1,4], MD, PhD

Andrea M. Burden[1], PhD

[1] Institute of Pharmaceutical Sciences, Department of Chemistry and Applied Biosciences, ETH Zurich, Zurich, Switzerland.

[2] Swiss Data Science Center, ETH Zurich and EPFL, Switzerland

[3] Institute of Machine Learning in the Computer Science Department at ETH Zurich

[4] Clinical Pharmacology and Toxicology, Department of General Internal Medicine, Inselspital, Bern University Hospital, University of Bern, Bern, Switzerland

### Corresponding Author

Prof. Dr. Andrea Burden

Institute of Pharmaceutical Sciences, Department of Chemistry and Applied Biosciences

ETH Zurich

Vladimir-Prelog-Weg 1-5/10

8093 Zurich

Switzerland

**Supplementary Table S1.** All chronic comorbidities selected for screening and whether they were included in the final analysis. In order to avoid convergence problems only comorbidities with a prevalence higher than 1% in the last period both overall and stratified by sex were included.

| Read Code | Description | Included | Count (%) |
|---|---|---|---|
| B | Cancer | Yes | 16981 (9.7) |
| G20 | High blood pressure | Yes | 11182 (6.4) |
| N05 | Osteoarthritis | Yes | 9210 (5.3) |
| F46 | Senile cataract | Yes | 7890 (4.5) |
| G57 | Atrial fibrillation | Yes | 7167 (4.1) |
| C32 | Pure hypercholesterolaemia | Yes | 6221 (3.5) |
| F59 | Deafness | Yes | 5442 (3.1) |
| J52 | Irritable bowel syndrome | Yes | 5517 (3.1) |
| E20 | Anxiety & other neurotic, stress related & somatoform disorders | Yes | 4830 (2.8) |
| J61 | Chronic liver disease | Yes | 4734 (2.7) |
| G58 | Congestive heart failure | Yes | 4058 (2.3) |
| C04 | Hypothyroidism | Yes | 3545 (2) |
| C38 | Obesity | Yes | 3283 (1.9) |
| N33 | Osteoporosis | Yes | 3381 (1.9) |
| G73 | Intermittent claudication | Yes | 2766 (1.6) |
| F45 | Primary open-angle glaucoma | Yes | 2410 (1.4) |
| H31 | Chronic bronchitis | Yes | 2297 (1.3) |
| N06 | Arthropathy | Yes | 2245 (1.3) |
| F36 | Hereditary and idiopathic peripheral neuropathy | Yes | 2038 (1.2) |
| M16 | Psoriasis or eczema | Yes | 2156 (1.2) |
| K05 | Chronic kidney disease | Yes | 1902 (1.1) |
| N11 | Cervical spondylosis | Yes | 2011 (1.1) |
| G33 | Angina pectoris | Yes | 1825 (1) |
| K20 | Prostatism | No | 1893 (1.1) |
| G34 | Ischaemic heart disease | No | 1161 (0.7) |
| E00 | Senile/presenile dementia | No | 684 (0.4) |
| H34 | Bronchiectasis | No | 651 (0.4) |
| N04 | Rheumatoid arthritis | No | 789 (0.4) |
| F12 | Parkinson’s disease | No | 590 (0.3) |
| F49 | Blindness & low vision | No | 532 (0.3) |
| H56 | Diffuse pulmonary fibrosis | No | 450 (0.3) |
| F25 | Epilepsy | No | 413 (0.2) |
| G67 | Cerebral atherosclerosis | No | 321 (0.2) |
| H32 | Emphysema | No | 371 (0.2) |
| H54 | Pulmonary oedema | No | 391 (0.2) |
| J41 | Ulcerative Colitis | No | 286 (0.2) |
| K11 | Hydronephrosis | No | 384 (0.2) |
| A70 | Viral Hepatitis | No | 100 (0.1) |
| D10 | Hereditary haemolytic anaemia | No | 155 (0.1) |
| E10 | Schizophrenia (and related non-organic psychosis) or bipolar disorder | No | 113 (0.1) |
| E2B1 | Chronic Depression | No | 160 (0.1) |
| F22 | Hemiplegia or paraplegia | No | 218 (0.1) |
| F24 | Hemiplegia or paraplegia | No | 205 (0.1) |
| G11 | Mitral stenosis | No | 97 (0.1) |
| J40 | Crohn's Disease | No | 136 (0.1) |
| F20 | Multiple sclerosis | No | 74 (0) |
| G21 | Hypertensive heart disease | No | 45 (0) |
| G22 | Hypertensive renal disease | No | 24 (0) |
| H35 | Extrinsic allergic alveolitis | No | 54 (0) |
| J13 | Peptic ulcer disease | No | 30 (0) |
| K03 | Nephritis and nephropathy | No | 31 (0) |
| N31 | Paget's disease of bone | No | 62 (0) |
| SC2 | Late effect - nervous system injury | No | 39 (0) |

**Supplementary Table S2.** Description of clusters after Bayesian nonparametric model.

| Cluster | Latent features | Read term | Count | N cluster | Count prop | Total disease | Total dis. Prop | O/E ratio |
|---|---|---|---|---|---|---|---|---|
| 1 | LF1 | High blood pressure | 8396 | 147816 | 5.7% | 10660 | 6.1% | 0.93 |
| 1 | LF1 | Pure hypercholesterolaemia | 4570 | 147816 | 3.1% | 5938 | 3.4% | 0.91 |
| 1 | LF1 | Chronic liver disease | 3437 | 147816 | 2.3% | 4505 | 2.6% | 0.91 |
| 1 | LF1 | Anxiety & other | 3101 | 147816 | 2.1% | 4115 | 2.3% | 0.89 |
| 1 | LF1 | Cancer | 11985 | 147816 | 8.1% | 15937 | 9.1% | 0.89 |
| 1 | LF1 | Irritable bowel syndrome | 3580 | 147816 | 2.4% | 5019 | 2.9% | 0.85 |
| 1 | LF1 | Deafness | 3721 | 147816 | 2.5% | 5261 | 3.0% | 0.84 |
| 1 | LF1 | Osteoarthritis | 6292 | 147816 | 4.3% | 8954 | 5.1% | 0.83 |
| 1 | LF1 | Senile cataract | 5139 | 147816 | 3.5% | 7709 | 4.4% | 0.79 |
| 1 | LF1 | Atrial fibrillation | 4299 | 147816 | 2.9% | 7000 | 4.0% | 0.73 |
| 1 | LF1 | Psoriasis or eczema | 0 | 147816 | 0.0% | 1975 | 1.1% | 0.00 |
| 1 | LF1 | Arthropathy | 0 | 147816 | 0.0% | 2168 | 1.2% | 0.00 |
| 1 | LF1 | Obesity | 0 | 147816 | 0.0% | 2879 | 1.6% | 0.00 |
| 1 | LF1 | Cervical spondylosis | 0 | 147816 | 0.0% | 1937 | 1.1% | 0.00 |
| 1 | LF1 | Primary open-angle glaucoma | 0 | 147816 | 0.0% | 2330 | 1.3% | 0.00 |
| 1 | LF1 | Chronic kidney disease | 0 | 147816 | 0.0% | 1869 | 1.1% | 0.00 |
| 1 | LF1 | Chronic bronchitis | 0 | 147816 | 0.0% | 2260 | 1.3% | 0.00 |
| 1 | LF1 | Angina pectoris | 0 | 147816 | 0.0% | 1773 | 1.0% | 0.00 |
| 1 | LF1 | Congestive heart failure | 0 | 147816 | 0.0% | 3962 | 2.3% | 0.00 |
| 1 | LF1 | Intermittent claudication | 0 | 147816 | 0.0% | 2653 | 1.5% | 0.00 |
| 1 | LF1 | Hypothyroidism | 0 | 147816 | 0.0% | 3260 | 1.9% | 0.00 |
| 1 | LF1 | Neuropathy | 0 | 147816 | 0.0% | 1964 | 1.1% | 0.00 |
| 1 | LF1 | Osteoporosis | 0 | 147816 | 0.0% | 3233 | 1.8% | 0.00 |

**Definitions:** Count, numbers of patients with that disease in that cluster; N cluster, total number of individuals within that cluster; Count prop, proportion of patients who have that disease within a cluster; Total disease, overall number of patients with that disease; Total dis. Prop, overall proportion that have that disease; O/E ratio, observed to expected ratio.

**Supplementary Table S2.** Description of clusters after Bayesian nonparametric model.

| Cluster | Latent features | Read term | Count | N cluster | Count prop | Total disease | Total dis. Prop | O/E ratio |
|---|---|---|---|---|---|---|---|---|
| 2 | LF1 LF2 | Congestive heart failure | 2711 | 2766 | 98.0% | 3962 | 2.3% | 43.39 |
| 2 | LF1 LF2 | Atrial fibrillation | 776 | 2766 | 28.1% | 7000 | 4.0% | 7.03 |
| 2 | LF1 LF2 | Senile cataract | 281 | 2766 | 10.2% | 7709 | 4.4% | 2.31 |
| 2 | LF1 LF2 | Deafness | 168 | 2766 | 6.1% | 5261 | 3.0% | 2.02 |
| 2 | LF1 LF2 | Irritable bowel syndrome | 141 | 2766 | 5.1% | 5019 | 2.9% | 1.78 |
| 2 | LF1 LF2 | Cancer | 395 | 2766 | 14.3% | 15937 | 9.1% | 1.57 |
| 2 | LF1 LF2 | Osteoarthritis | 183 | 2766 | 6.6% | 8954 | 5.1% | 1.30 |
| 2 | LF1 LF2 | Anxiety & other | 75 | 2766 | 2.7% | 4115 | 2.3% | 1.16 |
| 2 | LF1 LF2 | Chronic liver disease | 82 | 2766 | 3.0% | 4505 | 2.6% | 1.15 |
| 2 | LF1 LF2 | High blood pressure | 151 | 2766 | 5.5% | 10660 | 6.1% | 0.90 |
| 2 | LF1 LF2 | Pure hypercholesterolaemia | 77 | 2766 | 2.8% | 5938 | 3.4% | 0.82 |
| 2 | LF1 LF2 | Osteoporosis | 0 | 2766 | 0.0% | 3233 | 1.8% | 0.00 |
| 2 | LF1 LF2 | Obesity | 0 | 2766 | 0.0% | 2879 | 1.6% | 0.00 |
| 2 | LF1 LF2 | Cervical spondylosis | 0 | 2766 | 0.0% | 1937 | 1.1% | 0.00 |
| 2 | LF1 LF2 | Angina pectoris | 0 | 2766 | 0.0% | 1773 | 1.0% | 0.00 |
| 2 | LF1 LF2 | Arthropathy | 0 | 2766 | 0.0% | 2168 | 1.2% | 0.00 |
| 2 | LF1 LF2 | Intermittent claudication | 0 | 2766 | 0.0% | 2653 | 1.5% | 0.00 |
| 2 | LF1 LF2 | Psoriasis or eczema | 0 | 2766 | 0.0% | 1975 | 1.1% | 0.00 |
| 2 | LF1 LF2 | Neuropathy | 0 | 2766 | 0.0% | 1964 | 1.1% | 0.00 |
| 2 | LF1 LF2 | Hypothyroidism | 0 | 2766 | 0.0% | 3260 | 1.9% | 0.00 |
| 2 | LF1 LF2 | Chronic kidney disease | 0 | 2766 | 0.0% | 1869 | 1.1% | 0.00 |
| 2 | LF1 LF2 | Primary open-angle glaucoma | 0 | 2766 | 0.0% | 2330 | 1.3% | 0.00 |
| 2 | LF1 LF2 | Chronic bronchitis | 0 | 2766 | 0.0% | 2260 | 1.3% | 0.00 |

**Definitions:** Count, numbers of patients with that disease in that cluster; N cluster, total number of individuals within that cluster; Count prop, proportion of patients who have that disease within a cluster; Total disease, overall number of patients with that disease; Total dis. Prop, overall proportion that have that disease; O/E ratio, observed to expected ratio

**Supplementary Table S2. (continued)** Description of clusters after Bayesian nonparametric model.

| Cluster | Latent features | Read term | Count | N cluster | Count prop | Total disease | Total dis. Prop | O/E ratio |
|---|---|---|---|---|---|---|---|---|
| 3 | LF1 LF3 | Hypothyroidism | 2535 | 2572 | 98.6% | 3260 | 1.9% | 53.02 |
| 3 | LF1 LF3 | Irritable bowel syndrome | 118 | 2572 | 4.6% | 5019 | 2.9% | 1.60 |
| 3 | LF1 LF3 | Anxiety & other | 92 | 2572 | 3.6% | 4115 | 2.3% | 1.52 |
| 3 | LF1 LF3 | Senile cataract | 168 | 2572 | 6.5% | 7709 | 4.4% | 1.49 |
| 3 | LF1 LF3 | Pure hypercholesterolaemia | 129 | 2572 | 5.0% | 5938 | 3.4% | 1.48 |
| 3 | LF1 LF3 | Cancer | 342 | 2572 | 13.3% | 15937 | 9.1% | 1.46 |
| 3 | LF1 LF3 | Chronic liver disease | 95 | 2572 | 3.7% | 4505 | 2.6% | 1.44 |
| 3 | LF1 LF3 | Osteoarthritis | 184 | 2572 | 7.2% | 8954 | 5.1% | 1.40 |
| 3 | LF1 LF3 | High blood pressure | 213 | 2572 | 8.3% | 10660 | 6.1% | 1.36 |
| 3 | LF1 LF3 | Deafness | 98 | 2572 | 3.8% | 5261 | 3.0% | 1.27 |
| 3 | LF1 LF3 | Atrial fibrillation | 121 | 2572 | 4.7% | 7000 | 4.0% | 1.18 |
| 3 | LF1 LF3 | Congestive heart failure | 0 | 2572 | 0.0% | 3962 | 2.3% | 0.00 |
| 3 | LF1 LF3 | Neuropathy | 0 | 2572 | 0.0% | 1964 | 1.1% | 0.00 |
| 3 | LF1 LF3 | Obesity | 0 | 2572 | 0.0% | 2879 | 1.6% | 0.00 |
| 3 | LF1 LF3 | Primary open-angle glaucoma | 0 | 2572 | 0.0% | 2330 | 1.3% | 0.00 |
| 3 | LF1 LF3 | Angina pectoris | 0 | 2572 | 0.0% | 1773 | 1.0% | 0.00 |
| 3 | LF1 LF3 | Psoriasis or eczema | 0 | 2572 | 0.0% | 1975 | 1.1% | 0.00 |
| 3 | LF1 LF3 | Arthropathy | 0 | 2572 | 0.0% | 2168 | 1.2% | 0.00 |
| 3 | LF1 LF3 | Osteoporosis | 0 | 2572 | 0.0% | 3233 | 1.8% | 0.00 |
| 3 | LF1 LF3 | Chronic kidney disease | 0 | 2572 | 0.0% | 1869 | 1.1% | 0.00 |
| 3 | LF1 LF3 | Cervical spondylosis | 0 | 2572 | 0.0% | 1937 | 1.1% | 0.00 |
| 3 | LF1 LF3 | Intermittent claudication | 0 | 2572 | 0.0% | 2653 | 1.5% | 0.00 |
| 3 | LF1 LF3 | Chronic bronchitis | 0 | 2572 | 0.0% | 2260 | 1.3% | 0.00 |

**Definitions:** Count, numbers of patients with that disease in that cluster; N cluster, total number of individuals within that cluster; Count prop, proportion of patients who have that disease within a cluster; Total disease, overall number of patients with that disease; Total dis. Prop, overall proportion that have that disease; O/E ratio, observed to expected ratio.

**Supplementary Table S2. (continued)** Description of clusters after Bayesian nonparametric model.

| Cluster | Latent features | Read term | Count | N cluster | Count prop | Total disease | Total dis. Prop | O/E ratio |
|---|---|---|---|---|---|---|---|---|
| 4 | LF1 LF4 | Osteoporosis | 2323 | 2350 | 98.9% | 3233 | 1.8% | 53.62 |
| 4 | LF1 LF4 | Senile cataract | 269 | 2350 | 11.4% | 7709 | 4.4% | 2.60 |
| 4 | LF1 LF4 | Irritable bowel syndrome | 148 | 2350 | 6.3% | 5019 | 2.9% | 2.20 |
| 4 | LF1 LF4 | Deafness | 144 | 2350 | 6.1% | 5261 | 3.0% | 2.04 |
| 4 | LF1 LF4 | Osteoarthritis | 212 | 2350 | 9.0% | 8954 | 5.1% | 1.77 |
| 4 | LF1 LF4 | Atrial fibrillation | 162 | 2350 | 6.9% | 7000 | 4.0% | 1.73 |
| 4 | LF1 LF4 | Cancer | 347 | 2350 | 14.8% | 15937 | 9.1% | 1.62 |
| 4 | LF1 LF4 | Anxiety & other | 81 | 2350 | 3.4% | 4115 | 2.3% | 1.47 |
| 4 | LF1 LF4 | Chronic liver disease | 85 | 2350 | 3.6% | 4505 | 2.6% | 1.41 |
| 4 | LF1 LF4 | Pure hypercholesterolaemia | 90 | 2350 | 3.8% | 5938 | 3.4% | 1.13 |
| 4 | LF1 LF4 | High blood pressure | 157 | 2350 | 6.7% | 10660 | 6.1% | 1.10 |
| 4 | LF1 LF4 | Hypothyroidism | 0 | 2350 | 0.0% | 3260 | 1.9% | 0.00 |
| 4 | LF1 LF4 | Obesity | 0 | 2350 | 0.0% | 2879 | 1.6% | 0.00 |
| 4 | LF1 LF4 | Congestive heart failure | 0 | 2350 | 0.0% | 3962 | 2.3% | 0.00 |
| 4 | LF1 LF4 | Angina pectoris | 0 | 2350 | 0.0% | 1773 | 1.0% | 0.00 |
| 4 | LF1 LF4 | Intermittent claudication | 0 | 2350 | 0.0% | 2653 | 1.5% | 0.00 |
| 4 | LF1 LF4 | Neuropathy | 0 | 2350 | 0.0% | 1964 | 1.1% | 0.00 |
| 4 | LF1 LF4 | Chronic kidney disease | 0 | 2350 | 0.0% | 1869 | 1.1% | 0.00 |
| 4 | LF1 LF4 | Cervical spondylosis | 0 | 2350 | 0.0% | 1937 | 1.1% | 0.00 |
| 4 | LF1 LF4 | Arthropathy | 0 | 2350 | 0.0% | 2168 | 1.2% | 0.00 |
| 4 | LF1 LF4 | Psoriasis or eczema | 0 | 2350 | 0.0% | 1975 | 1.1% | 0.00 |
| 4 | LF1 LF4 | Chronic bronchitis | 0 | 2350 | 0.0% | 2260 | 1.3% | 0.00 |
| 4 | LF1 LF4 | Primary open-angle glaucoma | 0 | 2350 | 0.0% | 2330 | 1.3% | 0.00 |

**Definitions:** Count, numbers of patients with that disease in that cluster; N cluster, total number of individuals within that cluster; Count prop, proportion of patients who have that disease within a cluster; Total disease, overall number of patients with that disease; Total dis. Prop, overall proportion that have that disease; O/E ratio, observed to expected ratio.

**Supplementary Table S2. (continued)** Description of clusters after Bayesian nonparametric model.

| Cluster | Latent features | Read term | Count | N cluster | Count prop | Total disease | Total dis. Prop | O/E ratio |
|---|---|---|---|---|---|---|---|---|
| 5 | LF1 LF5 | Obesity | 2235 | 2252 | 99.2% | 2879 | 1.6% | 60.46 |
| 5 | LF1 LF5 | Pure hypercholesterolaemia | 185 | 2252 | 8.2% | 5938 | 3.4% | 2.43 |
| 5 | LF1 LF5 | Chronic liver disease | 121 | 2252 | 5.4% | 4505 | 2.6% | 2.09 |
| 5 | LF1 LF5 | High blood pressure | 276 | 2252 | 12.3% | 10660 | 6.1% | 2.02 |
| 5 | LF1 LF5 | Anxiety & other | 105 | 2252 | 4.7% | 4115 | 2.3% | 1.99 |
| 5 | LF1 LF5 | Irritable bowel syndrome | 101 | 2252 | 4.5% | 5019 | 2.9% | 1.57 |
| 5 | LF1 LF5 | Osteoarthritis | 179 | 2252 | 7.9% | 8954 | 5.1% | 1.56 |
| 5 | LF1 LF5 | Cancer | 262 | 2252 | 11.6% | 15937 | 9.1% | 1.28 |
| 5 | LF1 LF5 | Atrial fibrillation | 112 | 2252 | 5.0% | 7000 | 4.0% | 1.25 |
| 5 | LF1 LF5 | Deafness | 63 | 2252 | 2.8% | 5261 | 3.0% | 0.93 |
| 5 | LF1 LF5 | Senile cataract | 89 | 2252 | 4.0% | 7709 | 4.4% | 0.90 |
| 5 | LF1 LF5 | Hypothyroidism | 0 | 2252 | 0.0% | 3260 | 1.9% | 0.00 |
| 5 | LF1 LF5 | Psoriasis or eczema | 0 | 2252 | 0.0% | 1975 | 1.1% | 0.00 |
| 5 | LF1 LF5 | Neuropathy | 0 | 2252 | 0.0% | 1964 | 1.1% | 0.00 |
| 5 | LF1 LF5 | Angina pectoris | 0 | 2252 | 0.0% | 1773 | 1.0% | 0.00 |
| 5 | LF1 LF5 | Chronic kidney disease | 0 | 2252 | 0.0% | 1869 | 1.1% | 0.00 |
| 5 | LF1 LF5 | Arthropathy | 0 | 2252 | 0.0% | 2168 | 1.2% | 0.00 |
| 5 | LF1 LF5 | Chronic bronchitis | 0 | 2252 | 0.0% | 2260 | 1.3% | 0.00 |
| 5 | LF1 LF5 | Cervical spondylosis | 0 | 2252 | 0.0% | 1937 | 1.1% | 0.00 |
| 5 | LF1 LF5 | Primary open-angle glaucoma | 0 | 2252 | 0.0% | 2330 | 1.3% | 0.00 |
| 5 | LF1 LF5 | Intermittent claudication | 0 | 2252 | 0.0% | 2653 | 1.5% | 0.00 |
| 5 | LF1 LF5 | Osteoporosis | 0 | 2252 | 0.0% | 3233 | 1.8% | 0.00 |
| 5 | LF1 LF5 | Congestive heart failure | 0 | 2252 | 0.0% | 3962 | 2.3% | 0.00 |

**Definitions:** Count, numbers of patients with that disease in that cluster; N cluster, total number of individuals within that cluster; Count prop, proportion of patients who have that disease within a cluster; Total disease, overall number of patients with that disease; Total dis. Prop, overall proportion that have that disease; O/E ratio, observed to expected ratio.

**Supplementary Table S2. (continued)** Description of clusters after Bayesian nonparametric model.

| Cluster | Latent features | Read term | Count | N cluster | Count prop | Total disease | Total dis. Prop | O/E ratio |
|---|---|---|---|---|---|---|---|---|
| 6 | LF1 LF6 | Intermittent claudication | 1855 | 1871 | 99.1% | 2653 | 1.5% | 65.54 |
| 6 | LF1 LF6 | Atrial fibrillation | 153 | 1871 | 8.2% | 7000 | 4.0% | 2.05 |
| 6 | LF1 LF6 | Senile cataract | 153 | 1871 | 8.2% | 7709 | 4.4% | 1.86 |
| 6 | LF1 LF6 | Deafness | 96 | 1871 | 5.1% | 5261 | 3.0% | 1.71 |
| 6 | LF1 LF6 | Irritable bowel syndrome | 84 | 1871 | 4.5% | 5019 | 2.9% | 1.57 |
| 6 | LF1 LF6 | Cancer | 260 | 1871 | 13.9% | 15937 | 9.1% | 1.53 |
| 6 | LF1 LF6 | High blood pressure | 159 | 1871 | 8.5% | 10660 | 6.1% | 1.40 |
| 6 | LF1 LF6 | Osteoarthritis | 129 | 1871 | 6.9% | 8954 | 5.1% | 1.35 |
| 6 | LF1 LF6 | Pure hypercholesterolaemia | 78 | 1871 | 4.2% | 5938 | 3.4% | 1.23 |
| 6 | LF1 LF6 | Anxiety & other | 49 | 1871 | 2.6% | 4115 | 2.3% | 1.12 |
| 6 | LF1 LF6 | Chronic liver disease | 33 | 1871 | 1.8% | 4505 | 2.6% | 0.69 |
| 6 | LF1 LF6 | Osteoporosis | 0 | 1871 | 0.0% | 3233 | 1.8% | 0.00 |
| 6 | LF1 LF6 | Hypothyroidism | 0 | 1871 | 0.0% | 3260 | 1.9% | 0.00 |
| 6 | LF1 LF6 | Congestive heart failure | 0 | 1871 | 0.0% | 3962 | 2.3% | 0.00 |
| 6 | LF1 LF6 | Arthropathy | 0 | 1871 | 0.0% | 2168 | 1.2% | 0.00 |
| 6 | LF1 LF6 | Primary open-angle glaucoma | 0 | 1871 | 0.0% | 2330 | 1.3% | 0.00 |
| 6 | LF1 LF6 | Angina pectoris | 0 | 1871 | 0.0% | 1773 | 1.0% | 0.00 |
| 6 | LF1 LF6 | Cervical spondylosis | 0 | 1871 | 0.0% | 1937 | 1.1% | 0.00 |
| 6 | LF1 LF6 | Psoriasis or eczema | 0 | 1871 | 0.0% | 1975 | 1.1% | 0.00 |
| 6 | LF1 LF6 | Chronic bronchitis | 0 | 1871 | 0.0% | 2260 | 1.3% | 0.00 |
| 6 | LF1 LF6 | Obesity | 0 | 1871 | 0.0% | 2879 | 1.6% | 0.00 |
| 6 | LF1 LF6 | Chronic kidney disease | 0 | 1871 | 0.0% | 1869 | 1.1% | 0.00 |
| 6 | LF1 LF6 | Neuropathy | 0 | 1871 | 0.0% | 1964 | 1.1% | 0.00 |

**Definitions:** Count, numbers of patients with that disease in that cluster; N cluster, total number of individuals within that cluster; Count prop, proportion of patients who have that disease within a cluster; Total disease, overall number of patients with that disease; Total dis. Prop, overall proportion that have that disease; O/E ratio, observed to expected ratio.

**Supplementary Table S2. (continued)** Description of clusters after Bayesian nonparametric model.

| Cluster | Latent features | Read term | Count | N cluster | Count prop | Total disease | Total dis. Prop | O/E ratio |
|---|---|---|---|---|---|---|---|---|
| 7 | LF1 LF7 | Primary open-angle glaucoma | 1789 | 1795 | 99.7% | 2330 | 1.3% | 75.02 |
| 7 | LF1 LF7 | Senile cataract | 289 | 1795 | 16.1% | 7709 | 4.4% | 3.66 |
| 7 | LF1 LF7 | Osteoarthritis | 143 | 1795 | 8.0% | 8954 | 5.1% | 1.56 |
| 7 | LF1 LF7 | Deafness | 81 | 1795 | 4.5% | 5261 | 3.0% | 1.50 |
| 7 | LF1 LF7 | Cancer | 234 | 1795 | 13.0% | 15937 | 9.1% | 1.43 |
| 7 | LF1 LF7 | High blood pressure | 144 | 1795 | 8.0% | 10660 | 6.1% | 1.32 |
| 7 | LF1 LF7 | Pure hypercholesterolaemia | 79 | 1795 | 4.4% | 5938 | 3.4% | 1.30 |
| 7 | LF1 LF7 | Anxiety & other | 52 | 1795 | 2.9% | 4115 | 2.3% | 1.23 |
| 7 | LF1 LF7 | Chronic liver disease | 55 | 1795 | 3.1% | 4505 | 2.6% | 1.19 |
| 7 | LF1 LF7 | Atrial fibrillation | 84 | 1795 | 4.7% | 7000 | 4.0% | 1.17 |
| 7 | LF1 LF7 | Irritable bowel syndrome | 60 | 1795 | 3.3% | 5019 | 2.9% | 1.17 |
| 7 | LF1 LF7 | Cervical spondylosis | 0 | 1795 | 0.0% | 1937 | 1.1% | 0.00 |
| 7 | LF1 LF7 | Arthropathy | 0 | 1795 | 0.0% | 2168 | 1.2% | 0.00 |
| 7 | LF1 LF7 | Psoriasis or eczema | 0 | 1795 | 0.0% | 1975 | 1.1% | 0.00 |
| 7 | LF1 LF7 | Angina pectoris | 0 | 1795 | 0.0% | 1773 | 1.0% | 0.00 |
| 7 | LF1 LF7 | Chronic bronchitis | 0 | 1795 | 0.0% | 2260 | 1.3% | 0.00 |
| 7 | LF1 LF7 | Osteoporosis | 0 | 1795 | 0.0% | 3233 | 1.8% | 0.00 |
| 7 | LF1 LF7 | Hypothyroidism | 0 | 1795 | 0.0% | 3260 | 1.9% | 0.00 |
| 7 | LF1 LF7 | Congestive heart failure | 0 | 1795 | 0.0% | 3962 | 2.3% | 0.00 |
| 7 | LF1 LF7 | Intermittent claudication | 0 | 1795 | 0.0% | 2653 | 1.5% | 0.00 |
| 7 | LF1 LF7 | Obesity | 0 | 1795 | 0.0% | 2879 | 1.6% | 0.00 |
| 7 | LF1 LF7 | Chronic kidney disease | 0 | 1795 | 0.0% | 1869 | 1.1% | 0.00 |
| 7 | LF1 LF7 | Neuropathy | 0 | 1795 | 0.0% | 1964 | 1.1% | 0.00 |

**Definitions:** Count, numbers of patients with that disease in that cluster; N cluster, total number of individuals within that cluster; Count prop, proportion of patients who have that disease within a cluster; Total disease, overall number of patients with that disease; Total dis. Prop, overall proportion that have that disease; O/E ratio, observed to expected ratio.

**Supplementary Table S2. (continued)** Description of clusters after Bayesian nonparametric model.

| Cluster | Latent features | Read term | Count | N cluster | Count prop | Total disease | Total dis. Prop | O/E ratio |
|---|---|---|---|---|---|---|---|---|
| 8 | LF1 LF9 | Arthropathy | 1531 | 1532 | 99.9% | 2168 | 1.2% | 80.84 |
| 8 | LF1 LF9 | Osteoarthritis | 290 | 1532 | 18.9% | 8954 | 5.1% | 3.71 |
| 8 | LF1 LF9 | Anxiety & other | 70 | 1532 | 4.6% | 4115 | 2.3% | 1.95 |
| 8 | LF1 LF9 | Deafness | 82 | 1532 | 5.4% | 5261 | 3.0% | 1.78 |
| 8 | LF1 LF9 | Irritable bowel syndrome | 74 | 1532 | 4.8% | 5019 | 2.9% | 1.69 |
| 8 | LF1 LF9 | Cancer | 206 | 1532 | 13.4% | 15937 | 9.1% | 1.48 |
| 8 | LF1 LF9 | Senile cataract | 97 | 1532 | 6.3% | 7709 | 4.4% | 1.44 |
| 8 | LF1 LF9 | Pure hypercholesterolaemia | 73 | 1532 | 4.8% | 5938 | 3.4% | 1.41 |
| 8 | LF1 LF9 | High blood pressure | 129 | 1532 | 8.4% | 10660 | 6.1% | 1.39 |
| 8 | LF1 LF9 | Chronic liver disease | 54 | 1532 | 3.5% | 4505 | 2.6% | 1.37 |
| 8 | LF1 LF9 | Atrial fibrillation | 73 | 1532 | 4.8% | 7000 | 4.0% | 1.19 |
| 8 | LF1 LF9 | Cervical spondylosis | 0 | 1532 | 0.0% | 1937 | 1.1% | 0.00 |
| 8 | LF1 LF9 | Osteoporosis | 0 | 1532 | 0.0% | 3233 | 1.8% | 0.00 |
| 8 | LF1 LF9 | Hypothyroidism | 0 | 1532 | 0.0% | 3260 | 1.9% | 0.00 |
| 8 | LF1 LF9 | Chronic kidney disease | 0 | 1532 | 0.0% | 1869 | 1.1% | 0.00 |
| 8 | LF1 LF9 | Chronic bronchitis | 0 | 1532 | 0.0% | 2260 | 1.3% | 0.00 |
| 8 | LF1 LF9 | Psoriasis or eczema | 0 | 1532 | 0.0% | 1975 | 1.1% | 0.00 |
| 8 | LF1 LF9 | Angina pectoris | 0 | 1532 | 0.0% | 1773 | 1.0% | 0.00 |
| 8 | LF1 LF9 | Intermittent claudication | 0 | 1532 | 0.0% | 2653 | 1.5% | 0.00 |
| 8 | LF1 LF9 | Congestive heart failure | 0 | 1532 | 0.0% | 3962 | 2.3% | 0.00 |
| 8 | LF1 LF9 | Neuropathy | 0 | 1532 | 0.0% | 1964 | 1.1% | 0.00 |
| 8 | LF1 LF9 | Obesity | 0 | 1532 | 0.0% | 2879 | 1.6% | 0.00 |
| 8 | LF1 LF9 | Primary open-angle glaucoma | 0 | 1532 | 0.0% | 2330 | 1.3% | 0.00 |

**Definitions:** Count, numbers of patients with that disease in that cluster; N cluster, total number of individuals within that cluster; Count prop, proportion of patients who have that disease within a cluster; Total disease, overall number of patients with that disease; Total dis. Prop, overall proportion that have that disease; O/E ratio, observed to expected ratio.

**Supplementary Table S2. (continued)** Description of clusters after Bayesian nonparametric model.

| Cluster | Latent features | Read term | Count | N cluster | Count prop | Total disease | Total dis. Prop | O/E ratio |
|---|---|---|---|---|---|---|---|---|
| 9 | LF1 LF8 | Chronic bronchitis | 1494 | 1496 | 99.9% | 2260 | 1.3% | 77.50 |
| 9 | LF1 LF8 | Deafness | 95 | 1496 | 6.4% | 5261 | 3.0% | 2.12 |
| 9 | LF1 LF8 | Senile cataract | 134 | 1496 | 9.0% | 7709 | 4.4% | 2.04 |
| 9 | LF1 LF8 | Atrial fibrillation | 121 | 1496 | 8.1% | 7000 | 4.0% | 2.03 |
| 9 | LF1 LF8 | Irritable bowel syndrome | 85 | 1496 | 5.7% | 5019 | 2.9% | 1.99 |
| 9 | LF1 LF8 | Cancer | 223 | 1496 | 14.9% | 15937 | 9.1% | 1.64 |
| 9 | LF1 LF8 | Anxiety & other | 52 | 1496 | 3.5% | 4115 | 2.3% | 1.48 |
| 9 | LF1 LF8 | Osteoarthritis | 112 | 1496 | 7.5% | 8954 | 5.1% | 1.47 |
| 9 | LF1 LF8 | Chronic liver disease | 38 | 1496 | 2.5% | 4505 | 2.6% | 0.99 |
| 9 | LF1 LF8 | Pure hypercholesterolaemia | 47 | 1496 | 3.1% | 5938 | 3.4% | 0.93 |
| 9 | LF1 LF8 | High blood pressure | 75 | 1496 | 5.0% | 10660 | 6.1% | 0.82 |
| 9 | LF1 LF8 | Cervical spondylosis | 0 | 1496 | 0.0% | 1937 | 1.1% | 0.00 |
| 9 | LF1 LF8 | Chronic kidney disease | 0 | 1496 | 0.0% | 1869 | 1.1% | 0.00 |
| 9 | LF1 LF8 | Osteoporosis | 0 | 1496 | 0.0% | 3233 | 1.8% | 0.00 |
| 9 | LF1 LF8 | Congestive heart failure | 0 | 1496 | 0.0% | 3962 | 2.3% | 0.00 |
| 9 | LF1 LF8 | Arthropathy | 0 | 1496 | 0.0% | 2168 | 1.2% | 0.00 |
| 9 | LF1 LF8 | Psoriasis or eczema | 0 | 1496 | 0.0% | 1975 | 1.1% | 0.00 |
| 9 | LF1 LF8 | Angina pectoris | 0 | 1496 | 0.0% | 1773 | 1.0% | 0.00 |
| 9 | LF1 LF8 | Primary open-angle glaucoma | 0 | 1496 | 0.0% | 2330 | 1.3% | 0.00 |
| 9 | LF1 LF8 | Intermittent claudication | 0 | 1496 | 0.0% | 2653 | 1.5% | 0.00 |
| 9 | LF1 LF8 | Neuropathy | 0 | 1496 | 0.0% | 1964 | 1.1% | 0.00 |
| 9 | LF1 LF8 | Hypothyroidism | 0 | 1496 | 0.0% | 3260 | 1.9% | 0.00 |
| 9 | LF1 LF8 | Obesity | 0 | 1496 | 0.0% | 2879 | 1.6% | 0.00 |

**Definitions:** Count, numbers of patients with that disease in that cluster; N cluster, total number of individuals within that cluster; Count prop, proportion of patients who have that disease within a cluster; Total disease, overall number of patients with that disease; Total dis. Prop, overall proportion that have that disease; O/E ratio, observed to expected ratio.

**Supplementary Table S2. (continued)** Description of clusters after Bayesian nonparametric model.

| Cluster | Latent features | Read term | Count | N cluster | Count prop | Total disease | Total dis. Prop | O/E ratio |
|---|---|---|---|---|---|---|---|---|
| 10 | LF1 LF10 | Psoriasis or eczema | 1477 | 1481 | 99.7% | 1975 | 1.1% | 88.56 |
| 10 | LF1 LF10 | Osteoarthritis | 144 | 1481 | 9.7% | 8954 | 5.1% | 1.90 |
| 10 | LF1 LF10 | Chronic liver disease | 71 | 1481 | 4.8% | 4505 | 2.6% | 1.87 |
| 10 | LF1 LF10 | Anxiety & other | 56 | 1481 | 3.8% | 4115 | 2.3% | 1.61 |
| 10 | LF1 LF10 | High blood pressure | 139 | 1481 | 9.4% | 10660 | 6.1% | 1.54 |
| 10 | LF1 LF10 | Pure hypercholesterolaemia | 77 | 1481 | 5.2% | 5938 | 3.4% | 1.54 |
| 10 | LF1 LF10 | Irritable bowel syndrome | 63 | 1481 | 4.3% | 5019 | 2.9% | 1.49 |
| 10 | LF1 LF10 | Cancer | 185 | 1481 | 12.5% | 15937 | 9.1% | 1.37 |
| 10 | LF1 LF10 | Deafness | 52 | 1481 | 3.5% | 5261 | 3.0% | 1.17 |
| 10 | LF1 LF10 | Senile cataract | 75 | 1481 | 5.1% | 7709 | 4.4% | 1.15 |
| 10 | LF1 LF10 | Atrial fibrillation | 57 | 1481 | 3.8% | 7000 | 4.0% | 0.96 |
| 10 | LF1 LF10 | Intermittent claudication | 0 | 1481 | 0.0% | 2653 | 1.5% | 0.00 |
| 10 | LF1 LF10 | Obesity | 0 | 1481 | 0.0% | 2879 | 1.6% | 0.00 |
| 10 | LF1 LF10 | Congestive heart failure | 0 | 1481 | 0.0% | 3962 | 2.3% | 0.00 |
| 10 | LF1 LF10 | Angina pectoris | 0 | 1481 | 0.0% | 1773 | 1.0% | 0.00 |
| 10 | LF1 LF10 | Hypothyroidism | 0 | 1481 | 0.0% | 3260 | 1.9% | 0.00 |
| 10 | LF1 LF10 | Chronic kidney disease | 0 | 1481 | 0.0% | 1869 | 1.1% | 0.00 |
| 10 | LF1 LF10 | Neuropathy | 0 | 1481 | 0.0% | 1964 | 1.1% | 0.00 |
| 10 | LF1 LF10 | Osteoporosis | 0 | 1481 | 0.0% | 3233 | 1.8% | 0.00 |
| 10 | LF1 LF10 | Primary open-angle glaucoma | 0 | 1481 | 0.0% | 2330 | 1.3% | 0.00 |
| 10 | LF1 LF10 | Chronic bronchitis | 0 | 1481 | 0.0% | 2260 | 1.3% | 0.00 |
| 10 | LF1 LF10 | Cervical spondylosis | 0 | 1481 | 0.0% | 1937 | 1.1% | 0.00 |
| 10 | LF1 LF10 | Arthropathy | 0 | 1481 | 0.0% | 2168 | 1.2% | 0.00 |

**Definitions:** Count, numbers of patients with that disease in that cluster; N cluster, total number of individuals within that cluster; Count prop, proportion of patients who have that disease within a cluster; Total disease, overall number of patients with that disease; Total dis. Prop, overall proportion that have that disease; O/E ratio, observed to expected ratio.

**Supplementary Table S2. (continued)** Description of clusters after Bayesian nonparametric model.

| Cluster | Latent features | Read term | Count | N cluster | Count prop | Total disease | Total dis. Prop | O/E ratio |
|---|---|---|---|---|---|---|---|---|
| 11 | LF1 LF12 | Cervical spondylosis | 1369 | 1371 | 99.9% | 1937 | 1.1% | 90.41 |
| 11 | LF1 LF12 | Osteoarthritis | 204 | 1371 | 14.9% | 8954 | 5.1% | 2.91 |
| 11 | LF1 LF12 | Irritable bowel syndrome | 97 | 1371 | 7.1% | 5019 | 2.9% | 2.47 |
| 11 | LF1 LF12 | Chronic liver disease | 72 | 1371 | 5.3% | 4505 | 2.6% | 2.04 |
| 11 | LF1 LF12 | Deafness | 77 | 1371 | 5.6% | 5261 | 3.0% | 1.87 |
| 11 | LF1 LF12 | Senile cataract | 108 | 1371 | 7.9% | 7709 | 4.4% | 1.79 |
| 11 | LF1 LF12 | Anxiety & other | 51 | 1371 | 3.7% | 4115 | 2.3% | 1.59 |
| 11 | LF1 LF12 | Cancer | 196 | 1371 | 14.3% | 15937 | 9.1% | 1.57 |
| 11 | LF1 LF12 | Pure hypercholesterolaemia | 69 | 1371 | 5.0% | 5938 | 3.4% | 1.49 |
| 11 | LF1 LF12 | High blood pressure | 116 | 1371 | 8.5% | 10660 | 6.1% | 1.39 |
| 11 | LF1 LF12 | Atrial fibrillation | 76 | 1371 | 5.5% | 7000 | 4.0% | 1.39 |
| 11 | LF1 LF12 | Congestive heart failure | 0 | 1371 | 0.0% | 3962 | 2.3% | 0.00 |
| 11 | LF1 LF12 | Chronic bronchitis | 0 | 1371 | 0.0% | 2260 | 1.3% | 0.00 |
| 11 | LF1 LF12 | Angina pectoris | 0 | 1371 | 0.0% | 1773 | 1.0% | 0.00 |
| 11 | LF1 LF12 | Primary open-angle glaucoma | 0 | 1371 | 0.0% | 2330 | 1.3% | 0.00 |
| 11 | LF1 LF12 | Chronic kidney disease | 0 | 1371 | 0.0% | 1869 | 1.1% | 0.00 |
| 11 | LF1 LF12 | Psoriasis or eczema | 0 | 1371 | 0.0% | 1975 | 1.1% | 0.00 |
| 11 | LF1 LF12 | Obesity | 0 | 1371 | 0.0% | 2879 | 1.6% | 0.00 |
| 11 | LF1 LF12 | Arthropathy | 0 | 1371 | 0.0% | 2168 | 1.2% | 0.00 |
| 11 | LF1 LF12 | Intermittent claudication | 0 | 1371 | 0.0% | 2653 | 1.5% | 0.00 |
| 11 | LF1 LF12 | Osteoporosis | 0 | 1371 | 0.0% | 3233 | 1.8% | 0.00 |
| 11 | LF1 LF12 | Neuropathy | 0 | 1371 | 0.0% | 1964 | 1.1% | 0.00 |
| 11 | LF1 LF12 | Hypothyroidism | 0 | 1371 | 0.0% | 3260 | 1.9% | 0.00 |

**Definitions:** Count, numbers of patients with that disease in that cluster; N cluster, total number of individuals within that cluster; Count prop, proportion of patients who have that disease within a cluster; Total disease, overall number of patients with that disease; Total dis. Prop, overall proportion that have that disease; O/E ratio, observed to expected ratio.

**Supplementary Table S2. (continued)** Description of clusters after Bayesian nonparametric model.

| Cluster | Latent features | Read term | Count | N cluster | Count prop | Total disease | Total dis. Prop | O/E ratio |
|---|---|---|---|---|---|---|---|---|
| 12 | LF1 LF11 | Neuropathy | 1350 | 1350 | 100.0% | 1964 | 1.1% | 89.30 |
| 12 | LF1 LF11 | Chronic liver disease | 65 | 1350 | 4.8% | 4505 | 2.6% | 1.87 |
| 12 | LF1 LF11 | Irritable bowel syndrome | 70 | 1350 | 5.2% | 5019 | 2.9% | 1.81 |
| 12 | LF1 LF11 | Osteoarthritis | 116 | 1350 | 8.6% | 8954 | 5.1% | 1.68 |
| 12 | LF1 LF11 | Deafness | 65 | 1350 | 4.8% | 5261 | 3.0% | 1.61 |
| 12 | LF1 LF11 | Senile cataract | 95 | 1350 | 7.0% | 7709 | 4.4% | 1.60 |
| 12 | LF1 LF11 | High blood pressure | 120 | 1350 | 8.9% | 10660 | 6.1% | 1.46 |
| 12 | LF1 LF11 | Cancer | 176 | 1350 | 13.0% | 15937 | 9.1% | 1.43 |
| 12 | LF1 LF11 | Pure hypercholesterolaemia | 65 | 1350 | 4.8% | 5938 | 3.4% | 1.42 |
| 12 | LF1 LF11 | Anxiety & other | 42 | 1350 | 3.1% | 4115 | 2.3% | 1.33 |
| 12 | LF1 LF11 | Atrial fibrillation | 66 | 1350 | 4.9% | 7000 | 4.0% | 1.22 |
| 12 | LF1 LF11 | Angina pectoris | 0 | 1350 | 0.0% | 1773 | 1.0% | 0.00 |
| 12 | LF1 LF11 | Intermittent claudication | 0 | 1350 | 0.0% | 2653 | 1.5% | 0.00 |
| 12 | LF1 LF11 | Chronic kidney disease | 0 | 1350 | 0.0% | 1869 | 1.1% | 0.00 |
| 12 | LF1 LF11 | Primary open-angle glaucoma | 0 | 1350 | 0.0% | 2330 | 1.3% | 0.00 |
| 12 | LF1 LF11 | Congestive heart failure | 0 | 1350 | 0.0% | 3962 | 2.3% | 0.00 |
| 12 | LF1 LF11 | Chronic bronchitis | 0 | 1350 | 0.0% | 2260 | 1.3% | 0.00 |
| 12 | LF1 LF11 | Obesity | 0 | 1350 | 0.0% | 2879 | 1.6% | 0.00 |
| 12 | LF1 LF11 | Hypothyroidism | 0 | 1350 | 0.0% | 3260 | 1.9% | 0.00 |
| 12 | LF1 LF11 | Cervical spondylosis | 0 | 1350 | 0.0% | 1937 | 1.1% | 0.00 |
| 12 | LF1 LF11 | Psoriasis or eczema | 0 | 1350 | 0.0% | 1975 | 1.1% | 0.00 |
| 12 | LF1 LF11 | Arthropathy | 0 | 1350 | 0.0% | 2168 | 1.2% | 0.00 |
| 12 | LF1 LF11 | Osteoporosis | 0 | 1350 | 0.0% | 3233 | 1.8% | 0.00 |

**Definitions:** Count, numbers of patients with that disease in that cluster; N cluster, total number of individuals within that cluster; Count prop, proportion of patients who have that disease within a cluster; Total disease, overall number of patients with that disease; Total dis. Prop, overall proportion that have that disease; O/E ratio, observed to expected ratio.

**Supplementary Table S2. (continued)** Description of clusters after Bayesian nonparametric model.

| Cluster | Latent features | Read term | Count | N cluster | Count prop | Total disease | Total dis. Prop | O/E ratio |
|---|---|---|---|---|---|---|---|---|
| 13 | LF1 LF14 | Angina pectoris | 1253 | 1255 | 99.8% | 1773 | 1.0% | 98.76 |
| 13 | LF1 LF14 | Deafness | 80 | 1255 | 6.4% | 5261 | 3.0% | 2.13 |
| 13 | LF1 LF14 | Atrial fibrillation | 105 | 1255 | 8.4% | 7000 | 4.0% | 2.10 |
| 13 | LF1 LF14 | Irritable bowel syndrome | 72 | 1255 | 5.7% | 5019 | 2.9% | 2.00 |
| 13 | LF1 LF14 | Anxiety & other | 56 | 1255 | 4.5% | 4115 | 2.3% | 1.90 |
| 13 | LF1 LF14 | Pure hypercholesterolaemia | 76 | 1255 | 6.1% | 5938 | 3.4% | 1.79 |
| 13 | LF1 LF14 | Osteoarthritis | 113 | 1255 | 9.0% | 8954 | 5.1% | 1.76 |
| 13 | LF1 LF14 | Senile cataract | 86 | 1255 | 6.9% | 7709 | 4.4% | 1.56 |
| 13 | LF1 LF14 | Chronic liver disease | 48 | 1255 | 3.8% | 4505 | 2.6% | 1.49 |
| 13 | LF1 LF14 | Cancer | 151 | 1255 | 12.0% | 15937 | 9.1% | 1.32 |
| 13 | LF1 LF14 | High blood pressure | 99 | 1255 | 7.9% | 10660 | 6.1% | 1.30 |
| 13 | LF1 LF14 | Obesity | 0 | 1255 | 0.0% | 2879 | 1.6% | 0.00 |
| 13 | LF1 LF14 | Psoriasis or eczema | 0 | 1255 | 0.0% | 1975 | 1.1% | 0.00 |
| 13 | LF1 LF14 | Primary open-angle glaucoma | 0 | 1255 | 0.0% | 2330 | 1.3% | 0.00 |
| 13 | LF1 LF14 | Hypothyroidism | 0 | 1255 | 0.0% | 3260 | 1.9% | 0.00 |
| 13 | LF1 LF14 | Osteoporosis | 0 | 1255 | 0.0% | 3233 | 1.8% | 0.00 |
| 13 | LF1 LF14 | Chronic kidney disease | 0 | 1255 | 0.0% | 1869 | 1.1% | 0.00 |
| 13 | LF1 LF14 | Neuropathy | 0 | 1255 | 0.0% | 1964 | 1.1% | 0.00 |
| 13 | LF1 LF14 | Cervical spondylosis | 0 | 1255 | 0.0% | 1937 | 1.1% | 0.00 |
| 13 | LF1 LF14 | Intermittent claudication | 0 | 1255 | 0.0% | 2653 | 1.5% | 0.00 |
| 13 | LF1 LF14 | Arthropathy | 0 | 1255 | 0.0% | 2168 | 1.2% | 0.00 |
| 13 | LF1 LF14 | Chronic bronchitis | 0 | 1255 | 0.0% | 2260 | 1.3% | 0.00 |
| 13 | LF1 LF14 | Congestive heart failure | 0 | 1255 | 0.0% | 3962 | 2.3% | 0.00 |

**Definitions:** Count, numbers of patients with that disease in that cluster; N cluster, total number of individuals within that cluster; Count prop, proportion of patients who have that disease within a cluster; Total disease, overall number of patients with that disease; Total dis. Prop, overall proportion that have that disease; O/E ratio, observed to expected ratio.

**Supplementary Table S2. (continued)** Description of clusters after Bayesian nonparametric model.

| Cluster | Latent features | Read term | Count | N cluster | Count prop | Total disease | Total dis. Prop | O/E ratio |
|---|---|---|---|---|---|---|---|---|
| 14 | LF1 LF13 | Chronic kidney disease | 1227 | 1228 | 99.9% | 1869 | 1.1% | 93.76 |
| 14 | LF1 LF13 | Atrial fibrillation | 109 | 1228 | 8.9% | 7000 | 4.0% | 2.22 |
| 14 | LF1 LF13 | Deafness | 81 | 1228 | 6.6% | 5261 | 3.0% | 2.20 |
| 14 | LF1 LF13 | Senile cataract | 118 | 1228 | 9.6% | 7709 | 4.4% | 2.19 |
| 14 | LF1 LF13 | Cancer | 195 | 1228 | 15.9% | 15937 | 9.1% | 1.75 |
| 14 | LF1 LF13 | High blood pressure | 104 | 1228 | 8.5% | 10660 | 6.1% | 1.39 |
| 14 | LF1 LF13 | Irritable bowel syndrome | 46 | 1228 | 3.7% | 5019 | 2.9% | 1.31 |
| 14 | LF1 LF13 | Pure hypercholesterolaemia | 54 | 1228 | 4.4% | 5938 | 3.4% | 1.30 |
| 14 | LF1 LF13 | Osteoarthritis | 68 | 1228 | 5.5% | 8954 | 5.1% | 1.08 |
| 14 | LF1 LF13 | Anxiety & other | 31 | 1228 | 2.5% | 4115 | 2.3% | 1.08 |
| 14 | LF1 LF13 | Chronic liver disease | 33 | 1228 | 2.7% | 4505 | 2.6% | 1.05 |
| 14 | LF1 LF13 | Osteoporosis | 0 | 1228 | 0.0% | 3233 | 1.8% | 0.00 |
| 14 | LF1 LF13 | Angina pectoris | 0 | 1228 | 0.0% | 1773 | 1.0% | 0.00 |
| 14 | LF1 LF13 | Psoriasis or eczema | 0 | 1228 | 0.0% | 1975 | 1.1% | 0.00 |
| 14 | LF1 LF13 | Hypothyroidism | 0 | 1228 | 0.0% | 3260 | 1.9% | 0.00 |
| 14 | LF1 LF13 | Arthropathy | 0 | 1228 | 0.0% | 2168 | 1.2% | 0.00 |
| 14 | LF1 LF13 | Intermittent claudication | 0 | 1228 | 0.0% | 2653 | 1.5% | 0.00 |
| 14 | LF1 LF13 | Cervical spondylosis | 0 | 1228 | 0.0% | 1937 | 1.1% | 0.00 |
| 14 | LF1 LF13 | Primary open-angle glaucoma | 0 | 1228 | 0.0% | 2330 | 1.3% | 0.00 |
| 14 | LF1 LF13 | Chronic bronchitis | 0 | 1228 | 0.0% | 2260 | 1.3% | 0.00 |
| 14 | LF1 LF13 | Congestive heart failure | 0 | 1228 | 0.0% | 3962 | 2.3% | 0.00 |
| 14 | LF1 LF13 | Neuropathy | 0 | 1228 | 0.0% | 1964 | 1.1% | 0.00 |
| 14 | LF1 LF13 | Obesity | 0 | 1228 | 0.0% | 2879 | 1.6% | 0.00 |

**Definitions:** Count, numbers of patients with that disease in that cluster; N cluster, total number of individuals within that cluster; Count prop, proportion of patients who have that disease within a cluster; Total disease, overall number of patients with that disease; Total dis. Prop, overall proportion that have that disease; O/E ratio, observed to expected ratio.

**Supplementary Table S2. (continued)** Description of clusters after Bayesian nonparametric model.

| Cluster | Latent features | Read term | Count | N cluster | Count prop. | Total disease | Total dis. Prop. | O/E ratio |
|---|---|---|---|---|---|---|---|---|
| 15 | LF1 LF13 LF2 | Chronic kidney disease | 135 | 135 | 100.0% | 1869 | 1.1% | 93.84 |
| 15 | LF1 LF13 LF2 | Congestive heart failure | 135 | 135 | 100.0% | 3962 | 2.3% | 44.27 |
| 15 | LF1 LF13 LF2 | Atrial fibrillation | 45 | 135 | 33.3% | 7000 | 4.0% | 8.35 |
| 15 | LF1 LF13 LF2 | Senile cataract | 17 | 135 | 12.6% | 7709 | 4.4% | 2.86 |
| 15 | LF1 LF13 LF2 | Cancer | 26 | 135 | 19.3% | 15937 | 9.1% | 2.12 |
| 15 | LF1 LF13 LF2 | Pure hypercholesterolaemia | 9 | 135 | 6.7% | 5938 | 3.4% | 1.97 |
| 15 | LF1 LF13 LF2 | Osteoarthritis | 10 | 135 | 7.4% | 8954 | 5.1% | 1.45 |
| 15 | LF1 LF13 LF2 | Irritable bowel syndrome | 5 | 135 | 3.7% | 5019 | 2.9% | 1.29 |
| 15 | LF1 LF13 LF2 | Deafness | 5 | 135 | 3.7% | 5261 | 3.0% | 1.23 |
| 15 | LF1 LF13 LF2 | High blood pressure | 10 | 135 | 7.4% | 10660 | 6.1% | 1.22 |
| 15 | LF1 LF13 LF2 | Chronic liver disease | 3 | 135 | 2.2% | 4505 | 2.6% | 0.87 |
| 15 | LF1 LF13 LF2 | Anxiety & other | 2 | 135 | 1.5% | 4115 | 2.3% | 0.63 |
| 15 | LF1 LF13 LF2 | Osteoporosis | 0 | 135 | 0.0% | 3233 | 1.8% | 0.00 |
| 15 | LF1 LF13 LF2 | Arthropathy | 0 | 135 | 0.0% | 2168 | 1.2% | 0.00 |
| 15 | LF1 LF13 LF2 | Cervical spondylosis | 0 | 135 | 0.0% | 1937 | 1.1% | 0.00 |
| 15 | LF1 LF13 LF2 | Chronic bronchitis | 0 | 135 | 0.0% | 2260 | 1.3% | 0.00 |
| 15 | LF1 LF13 LF2 | Intermittent claudication | 0 | 135 | 0.0% | 2653 | 1.5% | 0.00 |
| 15 | LF1 LF13 LF2 | Angina pectoris | 0 | 135 | 0.0% | 1773 | 1.0% | 0.00 |
| 15 | LF1 LF13 LF2 | Psoriasis or eczema | 0 | 135 | 0.0% | 1975 | 1.1% | 0.00 |
| 15 | LF1 LF13 LF2 | Neuropathy | 0 | 135 | 0.0% | 1964 | 1.1% | 0.00 |
| 15 | LF1 LF13 LF2 | Primary open-angle glaucoma | 0 | 135 | 0.0% | 2330 | 1.3% | 0.00 |
| 15 | LF1 LF13 LF2 | Obesity | 0 | 135 | 0.0% | 2879 | 1.6% | 0.00 |
| 15 | LF1 LF13 LF2 | Hypothyroidism | 0 | 135 | 0.0% | 3260 | 1.9% | 0.00 |

**Definitions:** Count, numbers of patients with that disease in that cluster; N cluster, total number of individuals within that cluster; Count prop, proportion of patients who have that disease within a cluster; Total disease, overall number of patients with that disease; Total dis. Prop, overall proportion that have that disease; O/E ratio, observed to expected ratio.

**Supplementary Table S2. (continued)** Description of clusters after Bayesian nonparametric model.

| Cluster | Latent features | Read term | Count | N cluster | Count prop. | Total disease | Total dis. Prop. | O/E ratio |
|---|---|---|---|---|---|---|---|---|
| 16 | LF1 LF8 LF2 | Chronic bronchitis | 126 | 126 | 100.0% | 2260 | 1.3% | 77.60 |
| 16 | LF1 LF8 LF2 | Congestive heart failure | 125 | 126 | 99.2% | 3962 | 2.3% | 43.91 |
| 16 | LF1 LF8 LF2 | Atrial fibrillation | 49 | 126 | 38.9% | 7000 | 4.0% | 9.74 |
| 16 | LF1 LF8 LF2 | Senile cataract | 21 | 126 | 16.7% | 7709 | 4.4% | 3.79 |
| 16 | LF1 LF8 LF2 | Osteoarthritis | 16 | 126 | 12.7% | 8954 | 5.1% | 2.49 |
| 16 | LF1 LF8 LF2 | Cancer | 23 | 126 | 18.3% | 15937 | 9.1% | 2.01 |
| 16 | LF1 LF8 LF2 | Pure hypercholesterolaemia | 8 | 126 | 6.3% | 5938 | 3.4% | 1.88 |
| 16 | LF1 LF8 LF2 | Anxiety & other | 5 | 126 | 4.0% | 4115 | 2.3% | 1.69 |
| 16 | LF1 LF8 LF2 | Irritable bowel syndrome | 5 | 126 | 4.0% | 5019 | 2.9% | 1.39 |
| 16 | LF1 LF8 LF2 | Deafness | 4 | 126 | 3.2% | 5261 | 3.0% | 1.06 |
| 16 | LF1 LF8 LF2 | Chronic liver disease | 3 | 126 | 2.4% | 4505 | 2.6% | 0.93 |
| 16 | LF1 LF8 LF2 | High blood pressure | 3 | 126 | 2.4% | 10660 | 6.1% | 0.39 |
| 16 | LF1 LF8 LF2 | Cervical spondylosis | 0 | 126 | 0.0% | 1937 | 1.1% | 0.00 |
| 16 | LF1 LF8 LF2 | Osteoporosis | 0 | 126 | 0.0% | 3233 | 1.8% | 0.00 |
| 16 | LF1 LF8 LF2 | Arthropathy | 0 | 126 | 0.0% | 2168 | 1.2% | 0.00 |
| 16 | LF1 LF8 LF2 | Primary open-angle glaucoma | 0 | 126 | 0.0% | 2330 | 1.3% | 0.00 |
| 16 | LF1 LF8 LF2 | Psoriasis or eczema | 0 | 126 | 0.0% | 1975 | 1.1% | 0.00 |
| 16 | LF1 LF8 LF2 | Angina pectoris | 0 | 126 | 0.0% | 1773 | 1.0% | 0.00 |
| 16 | LF1 LF8 LF2 | Intermittent claudication | 0 | 126 | 0.0% | 2653 | 1.5% | 0.00 |
| 16 | LF1 LF8 LF2 | Hypothyroidism | 0 | 126 | 0.0% | 3260 | 1.9% | 0.00 |
| 16 | LF1 LF8 LF2 | Chronic kidney disease | 0 | 126 | 0.0% | 1869 | 1.1% | 0.00 |
| 16 | LF1 LF8 LF2 | Obesity | 0 | 126 | 0.0% | 2879 | 1.6% | 0.00 |
| 16 | LF1 LF8 LF2 | Neuropathy | 0 | 126 | 0.0% | 1964 | 1.1% | 0.00 |

**Definitions:** Count, numbers of patients with that disease in that cluster; N cluster, total number of individuals within that cluster; Count prop, proportion of patients who have that disease within a cluster; Total disease, overall number of patients with that disease; Total dis. Prop, overall proportion that have that disease; O/E ratio, observed to expected ratio.

**Supplementary Table S2. (continued)** Description of clusters after Bayesian nonparametric model.

| Cluster | Latent features | Read term | Count | N cluster | Count prop. | Total disease | Total dis. Prop. | O/E ratio |
|---|---|---|---|---|---|---|---|---|
| 17 | LF1 V4 LF2 | Osteoporosis | 117 | 117 | 100.0% | 3233 | 1.8% | 54.25 |
| 17 | LF1 V4 LF2 | Congestive heart failure | 117 | 117 | 100.0% | 3962 | 2.3% | 44.27 |
| 17 | LF1 V4 LF2 | Atrial fibrillation | 29 | 117 | 24.8% | 7000 | 4.0% | 6.21 |
| 17 | LF1 V4 LF2 | Cancer | 30 | 117 | 25.6% | 15937 | 9.1% | 2.82 |
| 17 | LF1 V4 LF2 | Irritable bowel syndrome | 9 | 117 | 7.7% | 5019 | 2.9% | 2.69 |
| 17 | LF1 V4 LF2 | Senile cataract | 13 | 117 | 11.1% | 7709 | 4.4% | 2.53 |
| 17 | LF1 V4 LF2 | Anxiety & other | 5 | 117 | 4.3% | 4115 | 2.3% | 1.82 |
| 17 | LF1 V4 LF2 | Deafness | 5 | 117 | 4.3% | 5261 | 3.0% | 1.42 |
| 17 | LF1 V4 LF2 | Osteoarthritis | 8 | 117 | 6.8% | 8954 | 5.1% | 1.34 |
| 17 | LF1 V4 LF2 | Pure hypercholesterolaemia | 3 | 117 | 2.6% | 5938 | 3.4% | 0.76 |
| 17 | LF1 V4 LF2 | High blood pressure | 5 | 117 | 4.3% | 10660 | 6.1% | 0.70 |
| 17 | LF1 V4 LF2 | Chronic liver disease | 2 | 117 | 1.7% | 4505 | 2.6% | 0.67 |
| 17 | LF1 V4 LF2 | Chronic bronchitis | 0 | 117 | 0.0% | 2260 | 1.3% | 0.00 |
| 17 | LF1 V4 LF2 | Angina pectoris | 0 | 117 | 0.0% | 1773 | 1.0% | 0.00 |
| 17 | LF1 V4 LF2 | Intermittent claudication | 0 | 117 | 0.0% | 2653 | 1.5% | 0.00 |
| 17 | LF1 V4 LF2 | Primary open-angle glaucoma | 0 | 117 | 0.0% | 2330 | 1.3% | 0.00 |
| 17 | LF1 V4 LF2 | Obesity | 0 | 117 | 0.0% | 2879 | 1.6% | 0.00 |
| 17 | LF1 V4 LF2 | Neuropathy | 0 | 117 | 0.0% | 1964 | 1.1% | 0.00 |
| 17 | LF1 V4 LF2 | Chronic kidney disease | 0 | 117 | 0.0% | 1869 | 1.1% | 0.00 |
| 17 | LF1 V4 LF2 | Hypothyroidism | 0 | 117 | 0.0% | 3260 | 1.9% | 0.00 |
| 17 | LF1 V4 LF2 | Cervical spondylosis | 0 | 117 | 0.0% | 1937 | 1.1% | 0.00 |
| 17 | LF1 V4 LF2 | Arthropathy | 0 | 117 | 0.0% | 2168 | 1.2% | 0.00 |
| 17 | LF1 V4 LF2 | Psoriasis or eczema | 0 | 117 | 0.0% | 1975 | 1.1% | 0.00 |

**Definitions:** Count, numbers of patients with that disease in that cluster; N cluster, total number of individuals within that cluster; Count prop, proportion of patients who have that disease within a cluster; Total disease, overall number of patients with that disease; Total dis. Prop, overall proportion that have that disease; O/E ratio, observed to expected ratio.

**Supplementary Table S2. (continued)** Description of clusters after Bayesian nonparametric model.

| **Cluster** | **Latent features** | **Read term** | **Count** | **N cluster** | **Count prop.** | **Total disease** | **Total dis. Prop.** | **O/E ratio** |
|---|---|---|---|---|---|---|---|---|
| 18 | LF1 V3 LF2 | Hypothyroidism | 103 | 103 | 100.0% | 3260 | 1.9% | 53.80 |
| 18 | LF1 V3 LF2 | Congestive heart failure | 103 | 103 | 100.0% | 3962 | 2.3% | 44.27 |
| 18 | LF1 V3 LF2 | Atrial fibrillation | 35 | 103 | 34.0% | 7000 | 4.0% | 8.51 |
| 18 | LF1 V3 LF2 | Deafness | 11 | 103 | 10.7% | 5261 | 3.0% | 3.56 |
| 18 | LF1 V3 LF2 | Chronic liver disease | 8 | 103 | 7.8% | 4505 | 2.6% | 3.02 |
| 18 | LF1 V3 LF2 | Senile cataract | 11 | 103 | 10.7% | 7709 | 4.4% | 2.43 |
| 18 | LF1 V3 LF2 | Cancer | 19 | 103 | 18.4% | 15937 | 9.1% | 2.03 |
| 18 | LF1 V3 LF2 | Irritable bowel syndrome | 5 | 103 | 4.9% | 5019 | 2.9% | 1.70 |
| 18 | LF1 V3 LF2 | Anxiety & other | 3 | 103 | 2.9% | 4115 | 2.3% | 1.24 |
| 18 | LF1 V3 LF2 | Osteoarthritis | 6 | 103 | 5.8% | 8954 | 5.1% | 1.14 |
| 18 | LF1 V3 LF2 | High blood pressure | 6 | 103 | 5.8% | 10660 | 6.1% | 0.96 |
| 18 | LF1 V3 LF2 | Pure hypercholesterolaemia | 3 | 103 | 2.9% | 5938 | 3.4% | 0.86 |
| 18 | LF1 V3 LF2 | Chronic kidney disease | 0 | 103 | 0.0% | 1869 | 1.1% | 0.00 |
| 18 | LF1 V3 LF2 | Psoriasis or eczema | 0 | 103 | 0.0% | 1975 | 1.1% | 0.00 |
| 18 | LF1 V3 LF2 | Neuropathy | 0 | 103 | 0.0% | 1964 | 1.1% | 0.00 |
| 18 | LF1 V3 LF2 | Chronic bronchitis | 0 | 103 | 0.0% | 2260 | 1.3% | 0.00 |
| 18 | LF1 V3 LF2 | Intermittent claudication | 0 | 103 | 0.0% | 2653 | 1.5% | 0.00 |
| 18 | LF1 V3 LF2 | Obesity | 0 | 103 | 0.0% | 2879 | 1.6% | 0.00 |
| 18 | LF1 V3 LF2 | Osteoporosis | 0 | 103 | 0.0% | 3233 | 1.8% | 0.00 |
| 18 | LF1 V3 LF2 | Primary open-angle glaucoma | 0 | 103 | 0.0% | 2330 | 1.3% | 0.00 |
| 18 | LF1 V3 LF2 | Angina pectoris | 0 | 103 | 0.0% | 1773 | 1.0% | 0.00 |
| 18 | LF1 V3 LF2 | Arthropathy | 0 | 103 | 0.0% | 2168 | 1.2% | 0.00 |
| 18 | LF1 V3 LF2 | Cervical spondylosis | 0 | 103 | 0.0% | 1937 | 1.1% | 0.00 |

**Definitions:** Count, numbers of patients with that disease in that cluster; N cluster, total number of individuals within that cluster; Count prop, proportion of patients who have that disease within a cluster; Total disease, overall number of patients with that disease; Total dis. Prop, overall proportion that have that disease; O/E ratio, observed to expected ratio.

**Supplementary Table S2. (continued)** Description of clusters after Bayesian nonparametric model.

| Cluster | Latent features | Read term | Count | N cluster | Count prop. | Total disease | Total dis. Prop. | O/E ratio |
|---|---|---|---|---|---|---|---|---|
| 19 | LF1 LF6 LF2 | Intermittent claudication | 101 | 101 | 100.0% | 2653 | 1.5% | 66.11 |
| 19 | LF1 LF6 LF2 | Congestive heart failure | 101 | 101 | 100.0% | 3962 | 2.3% | 44.27 |
| 19 | LF1 LF6 LF2 | Atrial fibrillation | 32 | 101 | 31.7% | 7000 | 4.0% | 7.94 |
| 19 | LF1 LF6 LF2 | Deafness | 10 | 101 | 9.9% | 5261 | 3.0% | 3.30 |
| 19 | LF1 LF6 LF2 | Senile cataract | 14 | 101 | 13.9% | 7709 | 4.4% | 3.15 |
| 19 | LF1 LF6 LF2 | Irritable bowel syndrome | 9 | 101 | 8.9% | 5019 | 2.9% | 3.11 |
| 19 | LF1 LF6 LF2 | Pure hypercholesterolaemia | 9 | 101 | 8.9% | 5938 | 3.4% | 2.63 |
| 19 | LF1 LF6 LF2 | Osteoarthritis | 10 | 101 | 9.9% | 8954 | 5.1% | 1.94 |
| 19 | LF1 LF6 LF2 | Cancer | 17 | 101 | 16.8% | 15937 | 9.1% | 1.85 |
| 19 | LF1 LF6 LF2 | Anxiety & other | 4 | 101 | 4.0% | 4115 | 2.3% | 1.69 |
| 19 | LF1 LF6 LF2 | High blood pressure | 9 | 101 | 8.9% | 10660 | 6.1% | 1.47 |
| 19 | LF1 LF6 LF2 | Chronic liver disease | 1 | 101 | 1.0% | 4505 | 2.6% | 0.39 |
| 19 | LF1 LF6 LF2 | Obesity | 0 | 101 | 0.0% | 2879 | 1.6% | 0.00 |
| 19 | LF1 LF6 LF2 | Hypothyroidism | 0 | 101 | 0.0% | 3260 | 1.9% | 0.00 |
| 19 | LF1 LF6 LF2 | Chronic bronchitis | 0 | 101 | 0.0% | 2260 | 1.3% | 0.00 |
| 19 | LF1 LF6 LF2 | Neuropathy | 0 | 101 | 0.0% | 1964 | 1.1% | 0.00 |
| 19 | LF1 LF6 LF2 | Psoriasis or eczema | 0 | 101 | 0.0% | 1975 | 1.1% | 0.00 |
| 19 | LF1 LF6 LF2 | Arthropathy | 0 | 101 | 0.0% | 2168 | 1.2% | 0.00 |
| 19 | LF1 LF6 LF2 | Cervical spondylosis | 0 | 101 | 0.0% | 1937 | 1.1% | 0.00 |
| 19 | LF1 LF6 LF2 | Angina pectoris | 0 | 101 | 0.0% | 1773 | 1.0% | 0.00 |
| 19 | LF1 LF6 LF2 | Chronic kidney disease | 0 | 101 | 0.0% | 1869 | 1.1% | 0.00 |
| 19 | LF1 LF6 LF2 | Primary open-angle glaucoma | 0 | 101 | 0.0% | 2330 | 1.3% | 0.00 |
| 19 | LF1 LF6 LF2 | Osteoporosis | 0 | 101 | 0.0% | 3233 | 1.8% | 0.00 |

**Definitions:** Count, numbers of patients with that disease in that cluster; N cluster, total number of individuals within that cluster; Count prop, proportion of patients who have that disease within a cluster; Total disease, overall number of patients with that disease; Total dis. Prop, overall proportion that have that disease; O/E ratio, observed to expected ratio.

**Supplementary Table S2. (continued)** Description of clusters after Bayesian nonparametric model.

| Cluster | Latent features | Read term | Count | N cluster | Count prop. | Total disease | Total dis. Prop. | O/E ratio |
|---|---|---|---|---|---|---|---|---|
| 20 | LF1 LF14 LF2 | Angina pectoris | 96 | 96 | 100.0% | 1773 | 1.0% | 98.92 |
| 20 | LF1 LF14 LF2 | Congestive heart failure | 96 | 96 | 100.0% | 3962 | 2.3% | 44.27 |
| 20 | LF1 LF14 LF2 | Atrial fibrillation | 25 | 96 | 26.0% | 7000 | 4.0% | 6.52 |
| 20 | LF1 LF14 LF2 | Senile cataract | 14 | 96 | 14.6% | 7709 | 4.4% | 3.32 |
| 20 | LF1 LF14 LF2 | Deafness | 8 | 96 | 8.3% | 5261 | 3.0% | 2.78 |
| 20 | LF1 LF14 LF2 | Irritable bowel syndrome | 6 | 96 | 6.3% | 5019 | 2.9% | 2.18 |
| 20 | LF1 LF14 LF2 | Cancer | 17 | 96 | 17.7% | 15937 | 9.1% | 1.95 |
| 20 | LF1 LF14 LF2 | Pure hypercholesterolaemia | 6 | 96 | 6.3% | 5938 | 3.4% | 1.85 |
| 20 | LF1 LF14 LF2 | Anxiety & other | 4 | 96 | 4.2% | 4115 | 2.3% | 1.78 |
| 20 | LF1 LF14 LF2 | Osteoarthritis | 7 | 96 | 7.3% | 8954 | 5.1% | 1.43 |
| 20 | LF1 LF14 LF2 | Chronic liver disease | 3 | 96 | 3.1% | 4505 | 2.6% | 1.22 |
| 20 | LF1 LF14 LF2 | High blood pressure | 5 | 96 | 5.2% | 10660 | 6.1% | 0.86 |
| 20 | LF1 LF14 LF2 | Obesity | 0 | 96 | 0.0% | 2879 | 1.6% | 0.00 |
| 20 | LF1 LF14 LF2 | Arthropathy | 0 | 96 | 0.0% | 2168 | 1.2% | 0.00 |
| 20 | LF1 LF14 LF2 | Hypothyroidism | 0 | 96 | 0.0% | 3260 | 1.9% | 0.00 |
| 20 | LF1 LF14 LF2 | Osteoporosis | 0 | 96 | 0.0% | 3233 | 1.8% | 0.00 |
| 20 | LF1 LF14 LF2 | Chronic kidney disease | 0 | 96 | 0.0% | 1869 | 1.1% | 0.00 |
| 20 | LF1 LF14 LF2 | Neuropathy | 0 | 96 | 0.0% | 1964 | 1.1% | 0.00 |
| 20 | LF1 LF14 LF2 | Chronic bronchitis | 0 | 96 | 0.0% | 2260 | 1.3% | 0.00 |
| 20 | LF1 LF14 LF2 | Intermittent claudication | 0 | 96 | 0.0% | 2653 | 1.5% | 0.00 |
| 20 | LF1 LF14 LF2 | Cervical spondylosis | 0 | 96 | 0.0% | 1937 | 1.1% | 0.00 |
| 20 | LF1 LF14 LF2 | Primary open-angle glaucoma | 0 | 96 | 0.0% | 2330 | 1.3% | 0.00 |
| 20 | LF1 LF14 LF2 | Psoriasis or eczema | 0 | 96 | 0.0% | 1975 | 1.1% | 0.00 |

**Definitions:** Count, numbers of patients with that disease in that cluster; N cluster, total number of individuals within that cluster; Count prop, proportion of patients who have that disease within a cluster; Total disease, overall number of patients with that disease; Total dis. Prop, overall proportion that have that disease; O/E ratio, observed to expected ratio.

**Supplementary Table S3.** Comparison of disease prevalence between the three main cardiovascular clusters.

| | | Cluster 2 (LF1 LF2) | | Cluster 15 (LF1 LF2 LF13) | | Cluster 16 (LF1 LF2 LF8) | | Proportions Comparison | |
|---|---|---|---|---|---|---|---|---|---|
| **Read term** | **Total dis. Prop** | **Count prop** | **O/E ratio** | **Count prop.** | **O/E ratio** | **Count prop.** | **O/E ratio** | **Cluster 15 vs Cluster 2** | **Cluster 16 vs Cluster 2** |
| Congestive heart failure | 2.3% | 98.0% | 43.39 | 100.0% | 44.27 | 99.2% | 43.91 | 1.02 | 1.01 |
| Atrial fibrillation | 4.0% | 28.1% | 7.03 | 33.3% | 8.35 | 38.9% | 9.74 | 1.19 | 1.39 |
| Senile cataract | 4.4% | 10.2% | 2.31 | 12.6% | 2.86 | 16.7% | 3.79 | 1.24 | 1.64 |
| Deafness | 3.0% | 6.1% | 2.02 | 3.7% | 1.23 | 3.2% | 1.06 | 0.61 | 0.52 |
| Irritable bowel syndrome | 2.9% | 5.1% | 1.78 | 3.7% | 1.29 | 4.0% | 1.39 | 0.73 | 0.78 |
| Cancer | 9.1% | 14.3% | 1.57 | 19.3% | 2.12 | 18.3% | 2.01 | 1.35 | 1.28 |
| Osteoarthritis | 5.1% | 6.6% | 1.30 | 7.4% | 1.45 | 12.7% | 2.49 | 1.12 | 1.92 |
| Anxiety & other | 2.3% | 2.7% | 1.16 | 1.5% | 0.63 | 4.0% | 1.69 | 0.55 | 1.46 |
| Chronic liver disease | 2.6% | 3.0% | 1.15 | 2.2% | 0.87 | 2.4% | 0.93 | 0.75 | 0.80 |
| High blood pressure | 6.1% | 5.5% | 0.90 | 7.4% | 1.22 | 2.4% | 0.39 | 1.36 | 0.44 |
| Pure hypercholesterolaemia | 3.4% | 2.8% | 0.82 | 6.7% | 1.97 | 6.3% | 1.88 | 2.39 | 2.28 |
| Osteoporosis | 1.8% | 0.0% | 0.00 | 0.0% | 0.00 | 0.0% | 0.00 | 0.00 | 0.00 |
| Obesity | 1.6% | 0.0% | 0.00 | 0.0% | 0.00 | 0.0% | 0.00 | 0.00 | 0.00 |
| Cervical spondylosis | 1.1% | 0.0% | 0.00 | 0.0% | 0.00 | 0.0% | 0.00 | 0.00 | 0.00 |
| Angina pectoris | 1.0% | 0.0% | 0.00 | 0.0% | 0.00 | 0.0% | 0.00 | 0.00 | 0.00 |
| Arthropathy | 1.2% | 0.0% | 0.00 | 0.0% | 0.00 | 0.0% | 0.00 | 0.00 | 0.00 |
| Intermittent claudication | 1.5% | 0.0% | 0.00 | 0.0% | 0.00 | 0.0% | 0.00 | 0.00 | 0.00 |
| Psoriasis or eczema | 1.1% | 0.0% | 0.00 | 0.0% | 0.00 | 0.0% | 0.00 | 0.00 | 0.00 |
| Neuropathy | 1.1% | 0.0% | 0.00 | 0.0% | 0.00 | 0.0% | 0.00 | 0.00 | 0.00 |
| Hypothyroidism | 1.9% | 0.0% | 0.00 | 0.0% | 0.00 | 0.0% | 0.00 | 0.00 | 0.00 |
| Chronic kidney disease | 1.1% | 0.0% | 0.00 | 100.0% | 93.84 | 0.0% | 0.00 | ∞ | 0.00 |
| Primary open-angle glaucoma | 1.3% | 0.0% | 0.00 | 0.0% | 0.00 | 0.0% | 0.00 | 0.00 | 0.00 |
| Chronic bronchitis | 1.3% | 0.0% | 0.00 | 0.0% | 0.00 | 100.0% | 77.60 | 0.00 | ∞ |

**Definitions:** Total dis. Prop, overall proportion that have that disease; Count prop, proportion of patients who have that disease within a cluster; O/E ratio, observed to expected ratio.

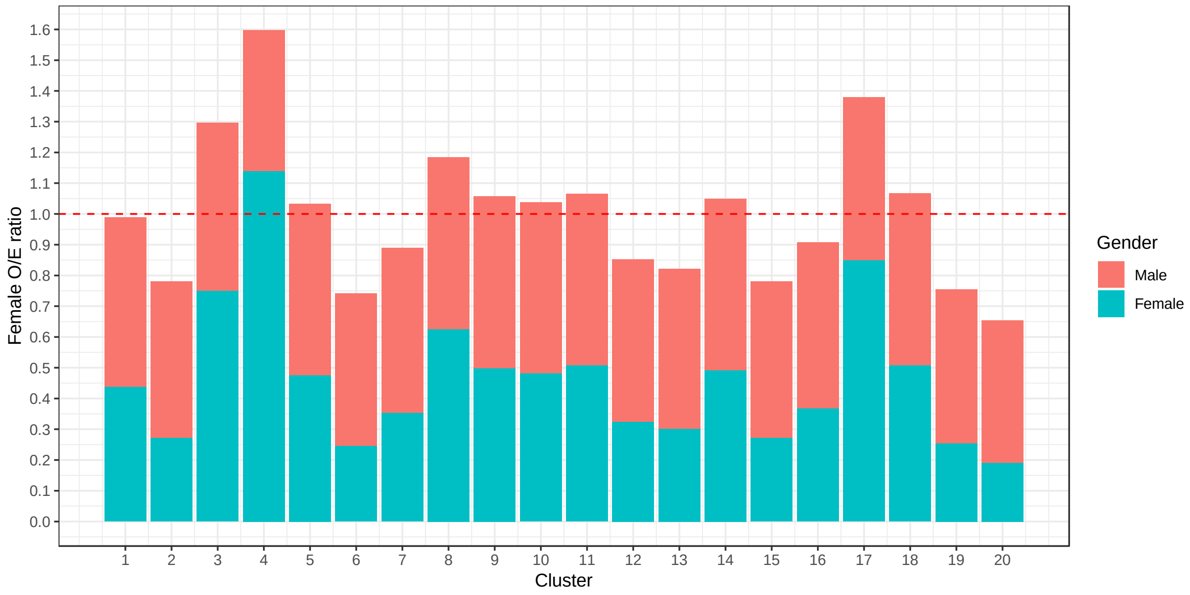


**Supplementary Figure S1.** O/E ratios for women for the first 20 clusters. The bars above O/E ratios 1.0 indicate that that specific cluster had a higher proportion of females than males. The colours represent the proportion of male and females in each cluster.

**Supplementary Figure S2.** Network representation of the transitions between first 20 clusters. Nodes represent clusters, edges indicate the transition between two given clusters, and the node size correspo to the logarithm of the size (number of patients in that node at any time). The nodes of clusters 15 to 20 had a low proportion of patients, thus a very small node size.

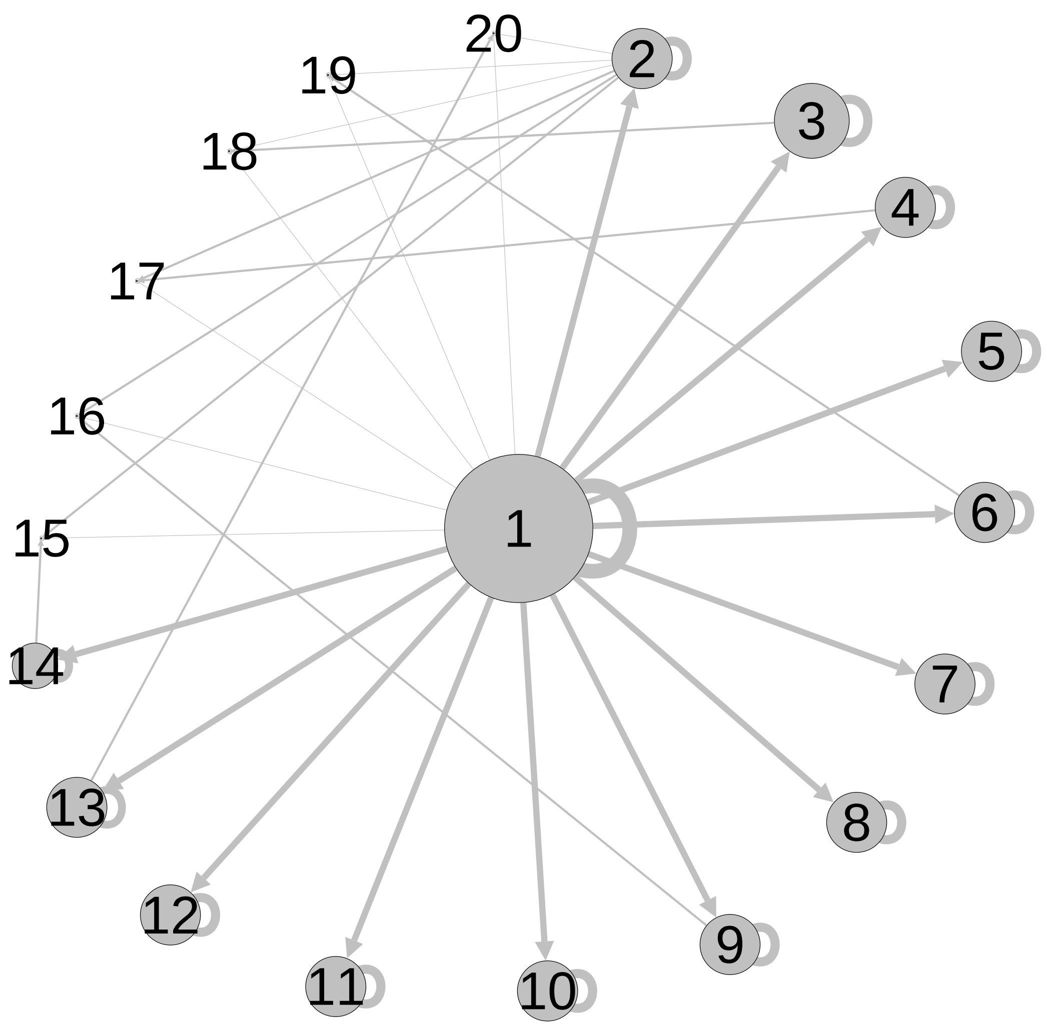
